# Dissecting Genetic and Environmental Determinants of Plasma Molecular Signatures and Their Link to Type 2 Diabetes Risk

**DOI:** 10.1101/2025.11.26.25341007

**Authors:** Magdalena Sevilla-González, Ningyuan Wang, Paul A. Hanson, Allison Bebo, Daniel Hitchcock, Sarah Hsu, Kenneth E. Westerman, Sara J. Cromer, Valene Garr Barry, Yonah Borns-Weil, Yixin Zhang, Ori Ben-Yossef, Chirag J. Patel, Nora Franceschini, Kent D. Taylor, Julian Ávila-Pacheco, Clary B. Clish, Robert E. Gertzen, Laura M. Raffield, Charles Kooperberg, Stephen S. Rich, Josée Dupuis, Jerome I. Rotter, Ching-Ti Liu, James B. Meigs, Alisa K. Manning, NHLBI Trans-Omics for Precision Medicine (TOPMed) Consortium

**Author notes:** **Corresponding Authors:** Alisa K, Manning, Ph.D. 100 Cambridge Street, Suite 1600, Clinical and Translational Epidemiology Unit Massachusetts General Hospital, Boston, MA, 02114, USA.; Magdalena Sevilla-Gonzalez, Ph.D. 100 Cambridge Street, Suite 1600, Clinical and Translational Epidemiology Unit Massachusetts General Hospital, Boston, MA, 02114, USA.

## Abstract

**Background:** Type 2 diabetes (T2D) is a heterogeneous disease shaped by both genetic, environmental, cultural, and socioeconomic factors, with well-documented disparities in incidence across populations. The molecular pathways underlying these disparities, however, remain poorly understood. Plasma metabolites and proteins integrate both genetic and environmental influences on type 2 diabetes (T2D) risk, providing insight into disease mechanisms. We aimed to quantify the variance in these molecular profiles explained by environmental and genetic ancestry domains and to apply causal inference approaches to identify environmentally and genetic ancestry influenced pathways contributing to T2D risk.

**Methods:** We analyzed plasma proteomic and metabolomic profiles from 3,360 MESA participants (51.6% female), and in 1,333 participants from the Women’s Health Initiative. To characterize the sources of variance in plasma proteomic and metabolomic profiles, we performed variance decomposition partitioning into four domains: biological (age, sex, BMI), genetic ancestry (principal components), lifestyle (smoking, alcohol intake, diet), and social determinants (self-reported race and ethnicity, income, education). To assess causal pathways towards T2D risk, we applied two-sample Mendelian Randomization to disentangle environmental and genetic contributors to T2D risk.

**Results:** The largest share of variance in proteomic and metabolomic profiles was explained by biological and lifestyle factors, while race and ethnicity and genetic ancestry accounted for smaller but non-redundant contributions. Genetic ancestry was primarily associated with lipid and apolipoprotein variation, whereas race and ethnicity and socioeconomic factors were associated with immune and inflammatory signatures. Environmentally influenced metabolites (e.g., diacylglycerols, phosphatidylethanolamines, lysophosphatidylcholines) and vascular–inflammatory proteins were consistently linked to higher T2D risk, while genetic ancestry influenced triglycerides and IGFBP3 reflected inherited risk pathways. Mediation analyses showed that selected lipids and proteins (e.g., IGFBP2, HGF, SSC4D) explained 10–25% of racial/ethnic disparities in T2D. Mendelian randomization identified causal roles for seven lipid species and IGFBP3 in T2D risk.

**Conclusions:** Our results reveal both genetic and non-genetic sources of variation in proteomic and metabolomic profiles, uncovering environmental and genetic pathways contributing to T2D risk. These findings advance precision medicine by identifying modifiable molecular mediators of disparities and potential causal targets for prevention.

## INTRODUCTION

Type 2 diabetes (T2D) is a heterogeneous disease arising from the interplay of genetic, environmental, cultural, and socioeconomic factors. By 2045, ∼11% of the global population (665 million individuals) is projected to have T2D.^1^ Disease disparities have been described in populations delineated by both genetic ancestry (a construct rooted in biology but with categorization based on geographical and historical social groupings) and by race and ethnicity, (a social construct rooted in systemic racism and structural inequities).^2^ While these population descriptors are related, they are distinct and reflect different sources of exposure/risk. The extent to which these overlapping but non-identical factors shape molecular profiles and contribute to health disparities in T2D remains largely unknown.

Metabolomics and proteomics, therefore, provide a unique lens to capture the convergence of genetic, environmental, and social influences on T2D pathophysiology. Plasma metabolites and proteins are powerful tools to identify biomarkers, elucidate mechanisms, and predict T2D risk.^3–11^ Because many directly participate in metabolic pathways, they also highlight potential therapeutic targets.^12^ However, most prior efforts have focused narrowly on genomic determinants or traditional risk factors, leaving the joint effects of social, environmental, and genetic domains on molecular variation underexplored. Insufficient attention has been given to disentangling the contributions of self-reported race and ethnicity—reflecting social and structural exposures—from genetic ancestry, which captures inherited biology. This gap is exacerbated by the underrepresentation of non-European populations in multi-omic datasets, limiting the generalizability of findings and constraining our understanding of causal pathways linking molecular signatures to T2D disparities. Additionally, clarifying the relative contributions of genetic and non-genetic factors is particularly important in contexts where population classification can be complex, as this may blur biological and social signals leading to misinterpretation of disease mechanisms or misguided prevention strategies.

In this study, we sought to disentangle the determinants of variability in plasma metabolites and proteins explained by biological, lifestyle, socioeconomic, race and ethnicity, and genetic ancestry domains. We further assessed the overlap and unique contributions of self-reported race and ethnicity versus genetic ancestry. Finally, we applied complementary causal inference approaches—mediation analysis and Mendelian randomization—to elucidate how environmental and genetic influences converge on pathways of T2D risk. The complete study design is presented in Figure 1.

**Figure 1.**
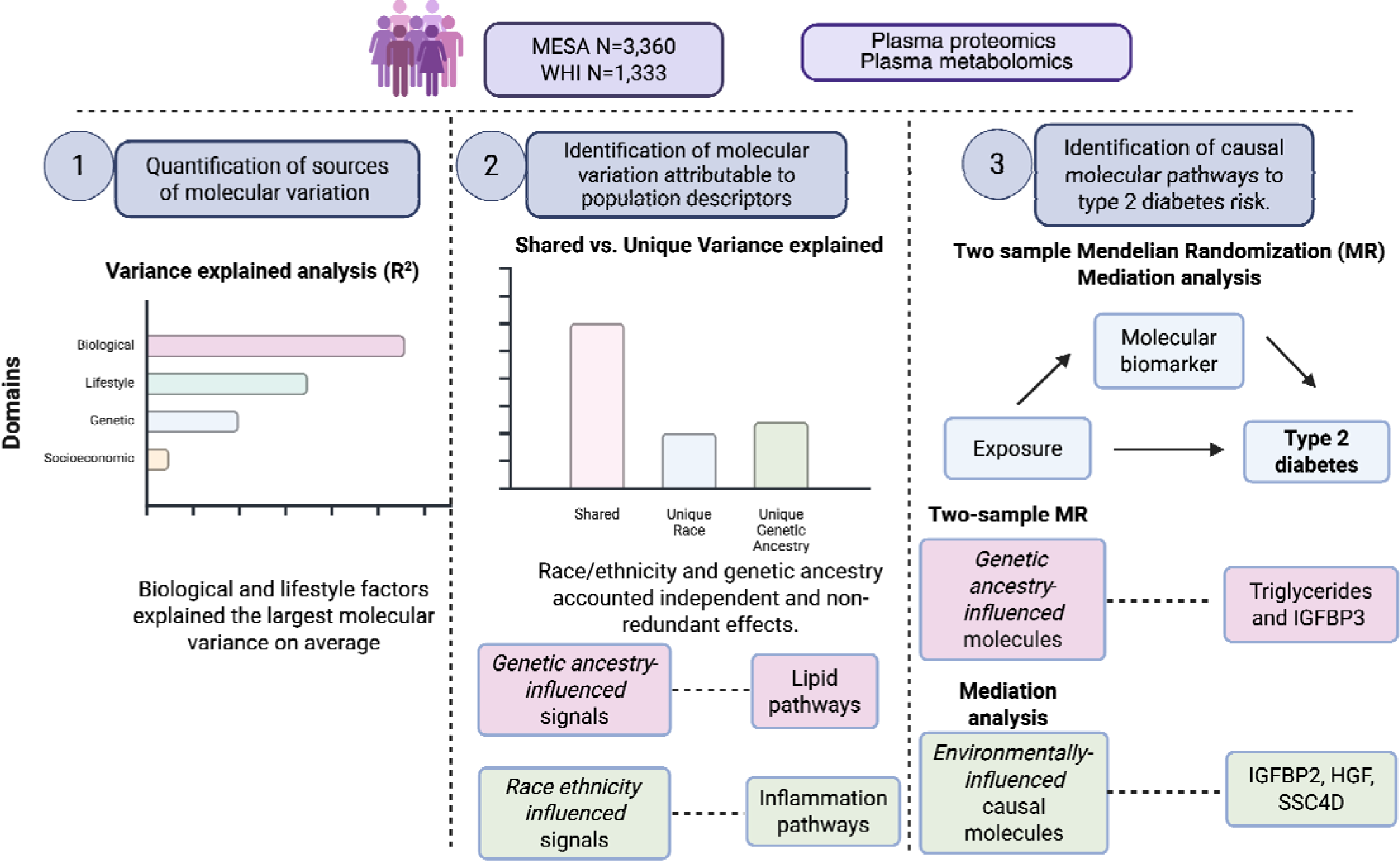
Study design. T2D Type 2 diabetes, MESA Multi-ethnic study of atherosclerosis, WHI Women’s Health Initiative. LC/MS Liquid Chromatography Mass Spectrometry. Figure created with BioRender.

## METHODS

### Population Description

Our study sample included participants from two cohorts of the Trans-Omics for Precision Medicine (TOPMed) program: the Multi-Ethnic Study of Atherosclerosis (MESA) and the Women’s Health Initiative (WHI). MESA is a population-based cohort designed to investigate the prevalence, correlates, and progression of subclinical cardiovascular disease in a diverse, multi-ethnic population. The WHI is a long-term national health study focused on the prevention of chronic diseases among postmenopausal women, including heart disease, breast and colorectal cancer, and osteoporosis. Detailed descriptions of each study design were published previously.^13,14^ Each study was approved by the Institutional Review Boards (IRBs) at all participating institutions, and all participants provided written informed consent at the start of the study.

### Metabolomic Data

Blood metabolite levels were quantified using liquid chromatography–mass spectrometry (LC-MS) at the Broad Institute Metabolomics Platform for both cohorts. Detailed descriptions of the metabolomics profiling protocols have been published previously^15^. In brief, plasma samples were analyzed using four complementary methods to maximize metabolome coverage: hydrophilic interaction chromatography and reversed-phase ultra-high performance liquid chromatography, each coupled with high-resolution mass spectrometry in both positive and negative ion modes. Targeted analysis of known metabolites was performed using TraceFinder (v3.3, Thermo Scientific), with metabolite identities confirmed through reference standards and pooled reference samples included in each analytical batch. High-resolution, non-targeted data were processed using Progenesis QI software (Nonlinear Dynamics) to detect peaks, align retention times, and integrate peak areas.

For this project, in MESA, we used fasting blood samples collected at Exam 1. In WHI, metabolites were measured at baseline as part of the Long-Life Study (LLS, 2012–2013), where fasting duration was recorded at the time of sample collection to account for potential metabolic variation.

Metabolomic datasets from the two cohorts were aligned using the Eclipse algorithm ^16^, which performs pairwise subalignments by scaling features across retention time, mass-to-charge ratio, and intensity residuals. Matched features are then aggregated into a directed graph, which is compressed and clustered to generate a consensus results table that was used in our analysis.

### Proteomic Data

In MESA, proteomic profiling was performed using the Olink Explore 3072 platform, which quantifies 2,941 antibody–oligonucleotide complexes corresponding to 2,923 unique UniProt identifiers. This high-throughput, multiplex technology is based on proximity extension assay (PEA), enabling sensitive and specific detection of low-abundance proteins in plasma.

In WHI, proteomic data were generated using six Olink Target panels (Cardiometabolic, Cardiovascular II, Cardiovascular III, Inflammation, Neurology, and Oncology III)^17^, comprising 552 proteins in total. Twelve proteins were measured across multiple panels; because their expression levels were highly correlated (average pairwise correlation = 0.925), we retained a single measurement for each duplicate, prioritizing proteins from cardiovascular and inflammation panels, yielding 540 proteins. All proteins included in the WHI dataset had overlapping UniProt identifiers with the MESA dataset.

For both cohorts, we used Normalized Protein eXpression (NPX) values provided by Olink for downstream analyses. NPX values are reported on a log2 scale and allow relative quantification and comparison of protein levels across samples.

### Covariate Assessment

We evaluated a comprehensive set of covariates spanning technical, biological, lifestyle, and social domains. Technical covariates included metabolomic batch (modeled as a categorical variable corresponding to the sample processing column), fasting hours at the time of blood draw, and site of recruitment. Biological covariates included age, sex (only in MESA), and genetic ancestry. Genetic ancestry was assessed using global ancestry proportions estimated with ADMIXTURE v1.3.1^18,19^, assuming four continental populations: African, American, European, and East Asian. Reference panels were assembled from the TOPMed dataset to ensure comparability with TOPMed study participants. The current TOPMed reference includes populations from both the 1000 Genomes Project (1KGP) and the Human Genome Diversity Panel (HGDP) (https://www.internationalgenome.org). Ancestry groups were defined as follows: African (AFR): Sub-Saharan African populations including HGDP (Bantu, Biaka, Mandenka, Mbuti, San, Yoruba) and 1KGP (Esan, Gambia, Luhya, Mende, Yoruba). European (EUR): HGDP (Bergamo, French, Sardinian, Tuscan) and 1KGP (Britain, Iberia, Toscani, Utah). American (AMR): HGDP (Colombian, Karitiana, Maya, Pima, Suruí) and 1KGP (Colombian, Peruvian). East Asian (EASIA): HGDP (Dai, Daur, Han, Hezhen, Japanese, Lahu, Miao, Mongolian, Naxi, Northern Han, Uygur, She, Tu, Tujia, Xibo, Yakut, Yi) and 1KGP (Dai, Han [Beijing and South], Japanese [Tokyo]). In addition, genetic principal components (PCs) were derived from genome-wide genotype data and included in subsequent analyses to account for population structure.

Self-reported race and ethnicity was defined as Non-Hispanic Black, non-Hispanic White, non-Hispanic Asian, Hispanic. Lifestyle covariates included smoking status (categorized as never, former, or current), alcohol intake (servings per day) and nutrient intake. Dietary variables were estimated from food frequency questionnaires and included energy intake (total kcals); % of total energy of macronutrients carbohydrates; fats; protein; grams of fiber; total sugars; saturated, monounsaturated, polyunsaturated, and trans fats; long-chain omega-3 fatty acids; omega-6 fatty acids; sodium; glycemic load; alcohol; vegetables; legumes; fruits; nuts; whole grains; red and processed meats; sugar-sweetened beverages; fruit juice; fish; and low-fat dairy. Socioeconomic covariates included education, and family income. Education was assessed using predefined cohort-specific categories ranging from no formal schooling to doctoral degree, while income was categorized from <$5,000/<$10,000 to ≥$100,000/≥$150,000 depending on cohort. In MESA, self-reported race and ethnicity, lifestyle covariates, and socioeconomic variables were assessed at the time of ‘omic measurements, whereas in WHI, these measures were obtained at enrollment (∼20 years prior to ‘omic profiling).

### Outcome Assessment

Incident T2D was defined as the first occurrence of any of the following criteria during follow-up after the ‘omics measurement: fasting glucose level >7.0 mmol/L (after ≥8 hours of fasting); initiation of diabetes treatment; hemoglobin A1c >6.5%; or physician diagnosis or self-reported diabetes.

### Statistical Analysis

#### Molecular Data Processing and Quality Control

Metabolite and protein data underwent a standardized preprocessing and quality control (QC) pipeline, following previously described approaches ^11,20^ and implemented through an open-source workflow available at https://github.com/manning-lab/metabolomics-qc. Features with zero variance across samples were excluded. Features and samples with >25% missingness were removed and remaining missing values were imputed using half of the minimum observed value for each feature under the assumption that missingness reflected concentrations below the limit of detection.^21^ To stabilize variance and improve normality, abundances were log□-transformed; Olink protein data were already log□-transformed by the provider. Outlier values were Winsorized to within ± 5 standard deviations of the mean to reduce the influence of extreme values while preserving overall data structure. All analyses were performed separately in each cohort to account for study-specific characteristics.

#### Multiple Testing Correction

To account for multiple testing in the ‘omic analyses, we estimated the number of effective independent tests by accounting for correlation among metabolites or proteins. This was performed using the meff function and the “liji” method from the poolr package in R^22^, which adjusts the significance threshold based on the underlying correlation structure.

#### Covariate Selection

We implemented a multi-step, data-driven framework to uncover the covariates that most strongly influence inter-individual variation in metabolomic and proteomic profiles. Metabolomic and proteomic principal components (PCs) were computed separately for each cohort. Each candidate covariate was evaluated for association with all ‘omic PCs using linear regression. Covariates were retained if they showed significant associations (P < 1×10) with at least one of the top ‘omic PCs that collectively captured the major sources of variation, as determined by eigenvalue decomposition.

#### Variance Decomposition and Covariate Contributions

We quantified the contribution of demographic, biological, technical, genetic, and social factors to variability in protein and metabolite abundances using linear regression-based variance decomposition. Covariates were grouped into different domains: technical (batch, fasting hours, recruitment site (MESA only), biological (age, sex, BMI), lifestyle (smoking, alcohol intake, dietary variables), genetic ancestry (first six genetic principal components), self-reported race and ethnicity, and social factors (income, education). The full model included all covariates. For each molecular feature, we estimated the variance explained (R^2^) by individual domains, the full model, and nested models excluding specific domains.

To assess domain-level contributions, we fit separate models for each domain, then compared these to the full model. Shared variance was calculated as the sum of block-level R^2^ minus the R^2^ of the full model, capturing overlap across domains. Next, domain-specific contributions were quantified as the mean unique R^2^ (%), representing the proportion of variance uniquely explained by each domain after accounting for all others. To evaluate statistical significance at the domain level, we applied ACAT method^23^ to combine p-values across features.

#### Classification of Molecular Features by Source of Variability

To classify molecular features according to their predominant source of variability, we computed feature-level genetic and environmental scores. For each molecule, a score was derived for both genetic ancestry and environmental influences by combining the effect size (R^2^) and statistical significance (−log_10_p) for each covariate block. The genetic score reflected the strength of association with genetic principal components, whereas the environmental score represented the average influence across socioeconomic status, race and ethnicity, and lifestyle factors. Molecules were then ranked independently within each omic type based on their computed scores. The top 30-ranked features with the highest genetic scores were classified as *genetic ancestry*-influenced molecules, while those with the highest environmental scores were classified as *environmentally influenced* molecules.

#### Race vs. Genetic Ancestry Decomposition

To disentangle the contributions of self-reported race and ethnicity and genetically inferred ancestry, we fit three models for each molecular biomarker: (A) race and ethnicity only, (B) genetic principal components (PCs) only, and (C) both race and PCs. Shared variance was defined as R^2^ (Model A) + R^2^ (Model B) - R^2^ (Model C), allowing us to determine the degree of overlap between race- and ancestry-related effects. Unique contributions were assessed using nested model comparisons, where reduced models excluding race or ancestry were compared with the full model to estimate ΔR^2^. Models were adjusted for technical, biological, lifestyle, and socioeconomic covariates. Additive models including both race and ancestry were used to evaluate joint explanatory power; differences between joint and summed contributions quantified shared variance. Finally, we applied ANOVA-based variance partitioning to directly compare variance explained by race/ethniciy and ancestry within the same population contrasts (e.g., Non-Hispanic Black vs. African ancestry, Hispanic vs. American ancestry, Non-Hispanic White vs. European).

#### Associations of Molecular Features with T2D Risk and Evaluation of Causal Pathways

To investigate the relationship between molecular features and the risk of T2D, we applied Cox proportional hazards models using time-to-event data among individuals free of T2D at baseline. Each model evaluated a single metabolite or protein as the exposure of interest and was adjusted for baseline technical, biological, lifestyle, and socioeconomic covariates. Hazard ratios (HRs) and 95% confidence intervals (CIs) were estimated for each molecular feature on the log scale.

We conducted a random-effects meta-analysis pooling cohort-specific estimates and standard errors. Proteins or metabolites were considered statistically significant if the meta-analysis yielded a *p-value < 0.05*. This threshold was chosen to balance sensitivity and specificity in the discovery stage, acknowledging the exploratory nature of these analyses. Subsequent causal inference and prioritization procedures applied more stringent multiple-testing corrections to minimize false positives.

To investigate potential causal molecular pathways underlying T2D risk, we applied complementary causal inference approaches tailored to the predominant source of molecular variability (environmental or genetic). For features with high genetic scores, we implemented Mendelian randomization (MR) to evaluate whether genetic ancestry influenced molecular variation was causally related to T2D risk. For molecular features with high environmental scores, we conducted mediation analyses to test whether environmentally influenced signatures mediated the association between self-reported race and ethnicity and incident T2D.

#### Mendelian Randomization Analyses

Mendelian randomization (MR) analyses were conducted to assess whether genetic ancestry influenced metabolites and proteins were causally related to T2D risk. Analyses were restricted to molecular features associated with T2D in the meta-analysis (*P < 0.05*) and, for metabolites, further limited to annotated metabolites with available Human Metabolome IDs (HMDBID) identifiers. Genetic instruments for metabolites were derived from multi-ancestry GWAS summary statistics from the TOPMed consortium ^24^, and for proteins from published *cis* pQTL data in UK Biobank.^25^ To avoid potential overfitting with our study cohorts (MESA and WHI), which were included in some large T2D GWAS meta-analyses, and to reflect the population diversity of our study cohorts we used multi-ancestry T2D GWAS summary statistics from the Million Veterans Program ^26^ as the outcome data source. Instrument selection applied allele frequency filtering (minor allele frequency > 0.01) and a distance-pruning algorithm, prioritizing the most significant variant at each locus and pruning all other variants within 1 million bp on either side of the variant to control for linkage disequilibrium. If two or more SNPs on the same chromosome were included in the final instrumental variable list, LD between the two or more SNPs was checked using an LDPair function from LDlink. All SNP pairs that had a D’ greater than 0.2 were pruned by taking the most significant SNP at that locus.

Two-sample MR analyses were conducted in R v4.3.1 using the TwoSampleMR package (v0.6.6). Three complementary MR methods were applied: inverse variance weighted (IVW), MR-Egger, and weighted median. Sensitivity analyses included heterogeneity tests using MR Egger and IVW, outlier tests using MR-PRESSO, and pleiotropy tests using MR Egger (P < 0.05). To account for multiple testing, Bonferroni correction was applied separately for the number of features tested in the mQTL and pQTL analyses, yielding corrected significance thresholds of P<0.001. An MR association was considered significant if the Bonferroni-corrected threshold was met in both the IVW and weighted median methods.

#### Mediation analysis

Mediation analyses focused on environmentally driven metabolites and proteins measured at baseline that were associated with incident T2D in MESA (p < 0.001). Analyses were restricted to MESA for two reasons: (i) the advanced age of WHI participants made diabetes diagnoses more likely to reflect age-related glycemic changes rather than true incident disease, and (ii) limited overlap in protein measurements between cohorts precluded harmonized analyses. MESA therefore provided a more reliable setting for evaluating causal pathways of environmentally influenced molecular signatures.

Analyses were conducted within the counterfactual (potential outcomes) framework using the cmest function from the CMAverse package.^27^ Self-reported race and ethnicity was modeled as a categorical exposure using pairwise comparisons. We focused on self-reported race and ethnicity as the exposure because it reflects structural and social determinants of health not fully captured by individual socioeconomic indicators such as income or education, which were instead included as covariates. T2D incidence was modeled with Cox proportional hazards regression, adjusting for a comprehensive set of covariates measured at baseline: age, sex, BMI, study site, smoking status, alcohol intake, fish, legumes, and red/processed meat intake, polyunsaturated and monounsaturated fat intake, total caloric intake, education, and income. A mediation effect was considered statistically significant if the proportion mediated was >10% and the corresponding *p-value* was *<1x 10⁻⁸*.

Mediation analyses were restricted to MESA for two reasons. First, the WHI cohort is substantially older, and T2D diagnoses in individuals >80 years are more likely to reflect age-related changes in glycemia rather than incident disease. Second, the overlap in protein measurements between WHI and MESA was limited, reducing the ability to perform comparable mediation analyses across cohorts. Together, these constraints made MESA the more reliable population for investigating mediation pathways.

## RESULTS

### Population

We leveraged 3,360 participants from the ancestrally diverse MESA cohort (51.6% women) for primary analyses and replicated findings in 1,333 participants from the WHI cohort. Participants in MESA had a mean age of 60 ± 9 years, whereas participants in WHI were considerably older, with a mean age of 81 ± 6 years. Additional demographic and clinical characteristics are provided in Supplementary Tables 1 and 2.

### Determinants of Molecular Variation

Metabolomic and proteomic principal components were significantly associated with technical, biological, genetic, lifestyle, and socioeconomic covariates (Supplementary Table 3; Supplementary Figures 1–4).

### Domain-Level Contributions to Molecular Variance

We next estimated the variance attributable to five covariate domains—biological (age, sex, BMI), lifestyle (smoking, alcohol, physical activity, diet), genetic ancestry, self-reported race and ethnicity, and socioeconomic status (SES)—using a unified multivariable framework. On average, these domains, in aggregate, explained ∼20% of metabolite variance and ∼12% of protein variance (Figure 2A). Contributions varied markedly across molecular biomarkers, with some dominated by biological or behavioral factors and others reflecting overlapping influences (Figure 2B–C).

**Figure 2.**
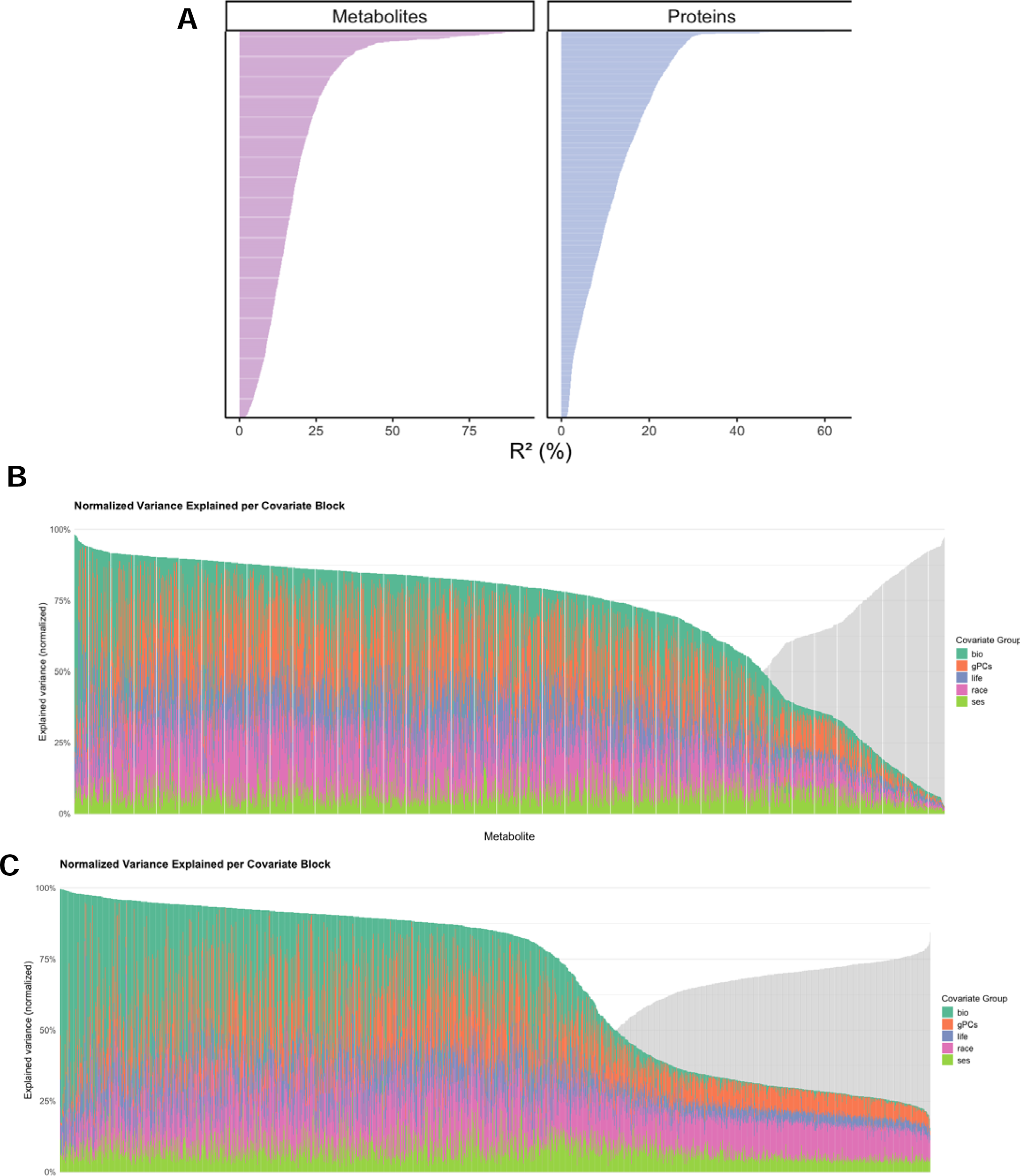
(A) **Total variance explained (R^2^) by the full linear regression model**—covariate domains include biological (age, sex, BMI), lifestyle (smoking, alcohol, physical activity, diet), genetic ancestry, self-reported race/ethnicity, and socioeconomic status (SES—across metabolites and proteins in MESA. **Normalized variance explained per covariate domain across individual metabolites (B) and proteins (C).** Normalized variance explained per covariate domain across individual metabolites (B) and proteins (C). For each molecule, R^2^ values were normalized to sum to 1 (100%), representing the relative contribution of each covariate domain. Bars are ordered by decreasing total variance explained.

Variance decomposition revealed that biological factors (age, sex, and BMI) accounted for the greatest unique variance in metabolites, on average, followed by lifestyle and SES, while genetic ancestry and self-reported race and ethnicity contributed smaller fractions, largely shared with other domains (Figure 3A). Results were similar for proteins, though the overall variance explained was lower. In both ‘omics layers, biological and lifestyle factors consistently emerged as dominant influences, whereas SES and race and ethnicity effects were smaller and more overlapping.

**Figure 3.**
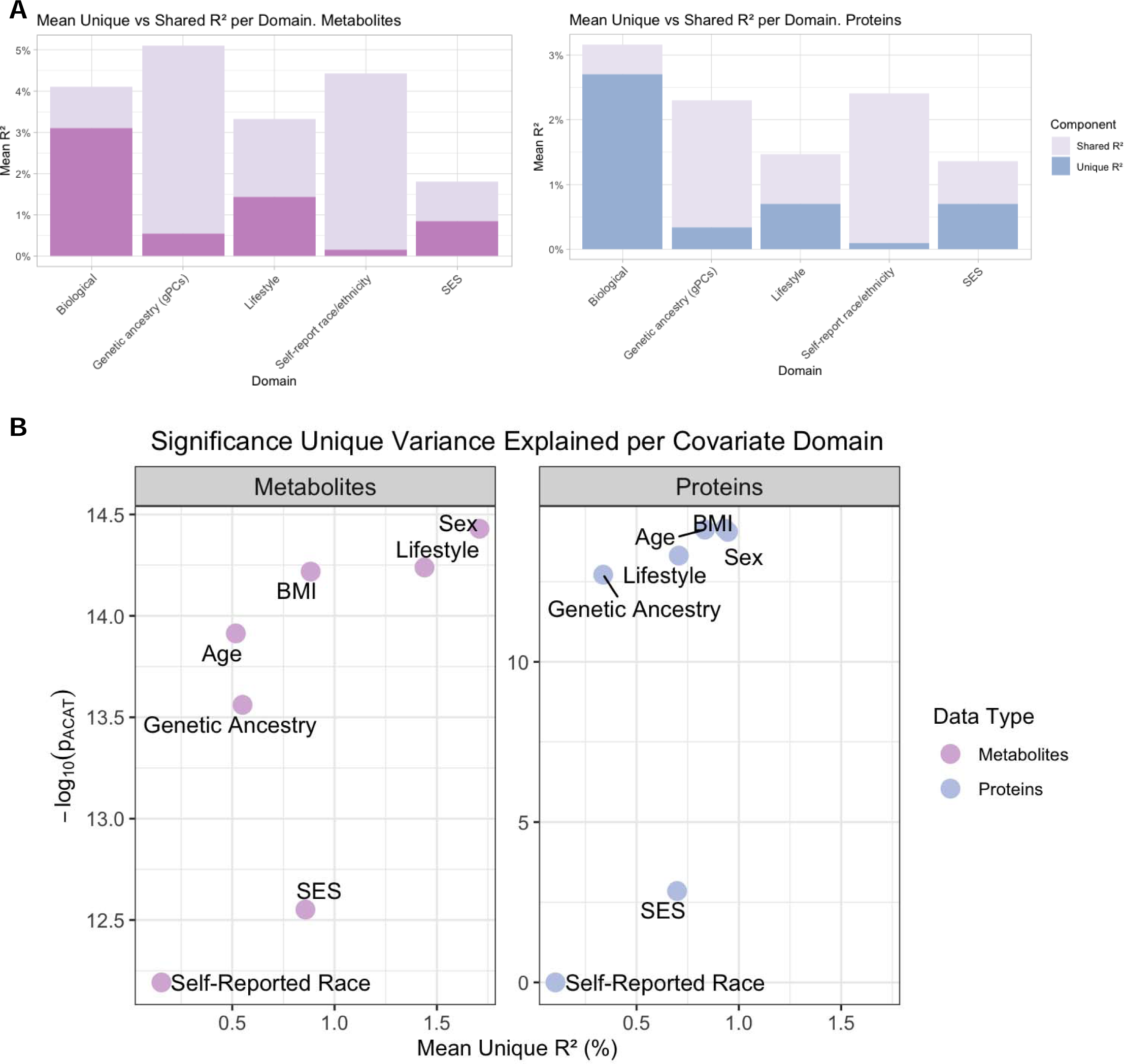
(A) **Mean variance explained (R^2^) in metabolites and proteins by covariate domains in MESA**, partitioned into unique and shared contributions. Bars represent the average proportion of variance in metabolite and protein levels explained by each domain (B). **Relationship between Statistical Significance and Unique Variance Explained.** The plot visualizes the contribution of various covariate domains to the variance in metabolites and proteins. The x-axis represents the mean unique variance explained (Unique R^2^, %) by each covariate domain. The y-axis shows the statistical significance of each covariate domain, quantified ACAT method.

As shown in Figure 3B, sex explained the greatest unique variance in metabolites on average, followed by lifestyle and BMI. Interestingly, SES contributed on average greater R^2^ than genetic ancestry and even age, albeit with lower p value on average. For proteins, BMI and age were the strongest contributors, with sex also significant, while lifestyle and SES contributed similarly on average. Importantly, both self-reported race and ethnicity and genetic ancestry made independent, statistically significant contributions when both included in our models, underscoring that these population-level constructs capture non-redundant aspects of molecular variability despite smaller magnitudes relative to biological and behavioral domains.

### Domain-Specific Influences on Individual Plasma Metabolites and Proteins

To highlight domain-specific molecular associations, we identified the top ten metabolites and proteins per domain, ranked by unique R² (Figure 4). BMI emerged as a strong contributor of metabolic variation, with robust associations for glutamic acid, multiple triglycerides, and proteins including FABP4, LEP, and IGFBP1. Lifestyle factors explained substantial variance in omega-3 fatty acids (EPA, DHA), cotinine, and proteins such as ALPP and CXCL17, providing reassuring, biologically consistent validation of lifestyle-driven molecular signatures.

**Figure 4.**
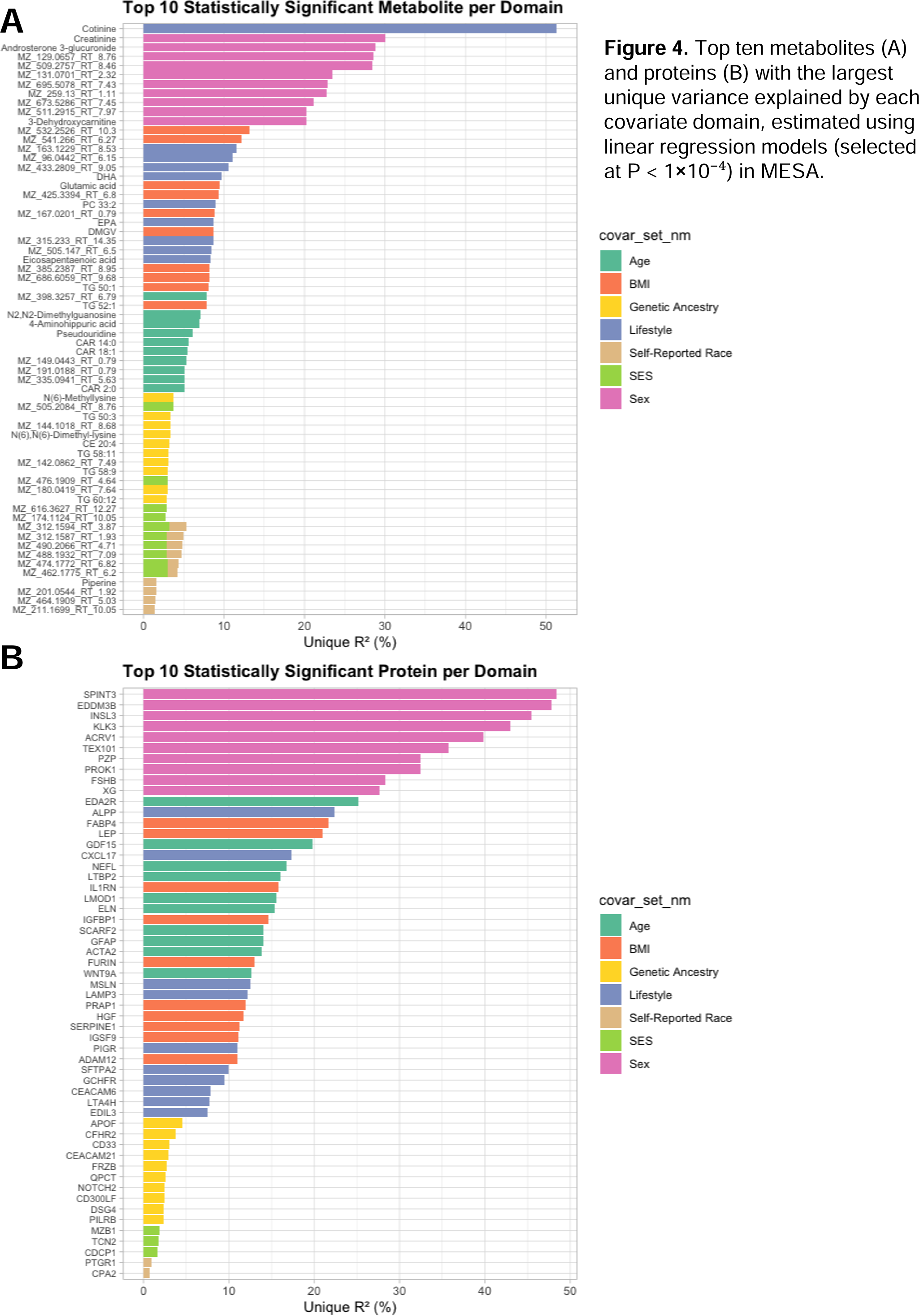
Top ten metabolites (A) and proteins (B) with the largest unique variance explained by each covariate domain, estimated using linear regression models (selected at P < 1×10^-4^) in MESA.

Although genetic ancestry and self-reported race and ethnicity explained smaller proportions of variance overall, they revealed highly specific associations. Genetic ancestry was linked to lipid-related molecules, including TG 50:3, TG 58:11, TG 60:12, N(6)-dimethyl lysine, and the minor apolipoprotein APOF—suggesting inherited genetic background shapes lipid subclass composition and apolipoprotein regulation. By contrast, self-reported race and ethnicity was enriched for inflammatory and immune-related proteins, including PTGR1 (a prostaglandin metabolism enzyme)^28^ and CPA2 (a pancreatic enzyme associated with inflammation).^29^ Notably, the top metabolites associated with race and ethnicity remained unannotated, highlighting novel, yet-to-be-characterized metabolic signatures. These results demonstrated strong consistency across the two cohorts, supporting the validity of our observations (Supplementary Figures 7–10).

### Race and ethnicity and Genetic Ancestry Explain Overlapping but Non-Redundant Molecular Variation

Race and ethnicity and genetic ancestry accounted for overlapping variance in molecular profiles but showed distinct biomarker-level effects: genetic ancestry influenced signals were enriched in lipid pathways, whereas race and ethnicity-related signatures predominantly reflected inflammation. We quantified the independent and shared contributions of self-reported race and ethnicity and genetic ancestry (gPCs) to plasma metabolomic and proteomic profiles. Across both ‘omics layers, much of the variance explained by these domains was shared; however, a subset of biomarkers exhibited distinct contributions from one construct over the other (Figure 5A–B). To disentangle the unique effects, we compared the mean variance explained (ΔR²) across all metabolites and proteins under different covariate models (Figure 5C). Genetic ancestry consistently accounted for a modest but significant proportion of variance after adjusting for other factors. Adding self-reported race and ethnicity to the model increased the mean variance explained, indicating that race and ethnicity captured non-overlapping sources of molecular variability beyond ancestry. By contrast, race and ethnicity alone explained less variance than ancestry, suggesting that genetic background contributes more strongly to global molecular differences, or alternatively, that broad categorical labels provide a coarser measure compared to the continuous spectrum captured by multiple genetic principal components. Comparing the summed variance of the two constructs with their combined model revealed a clear gap, highlighting substantial shared explanatory power. This pattern was observed for both metabolites and proteins in both cohorts, though effects were stronger in metabolites in MESA and in proteins in WHI (Supplementary Figure 10). Together, these results show that race and ethnicity and genetic ancestry are overlapping but non-redundant constructs, each capturing distinct sociocultural and biological dimensions of molecular variation.

**Figure 5.**
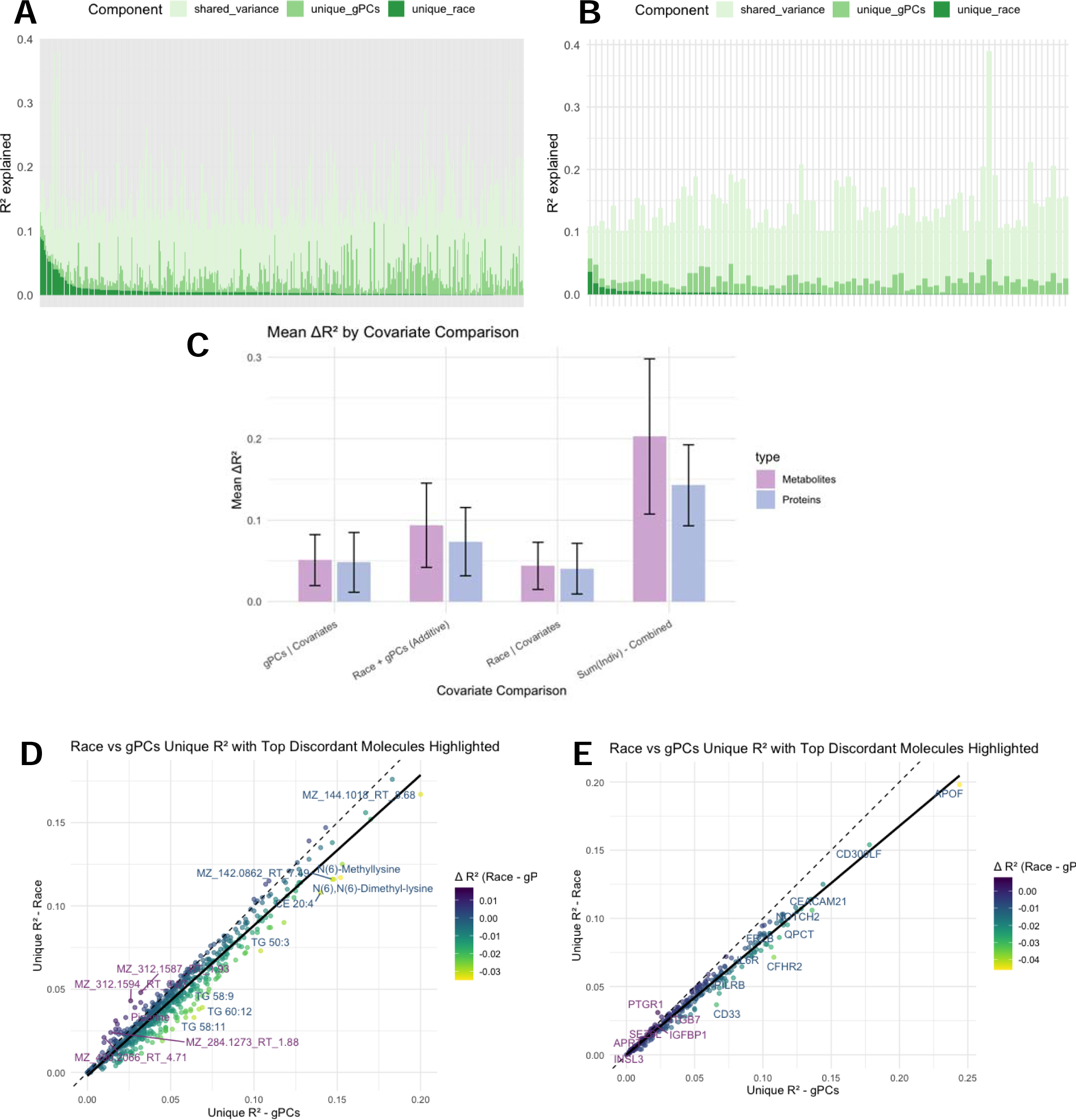
Independent and shared contributions of race/ethnicity and genetic ancestry to molecular variance in MESA. A, Variance explained (R²) for metabolites partitioned into shared variance, unique variance explained by genetic principal components (gPCs), and unique variance explained by self-reported race/ethnicity. B, Equivalent decomposition for proteins. Each bar represents one molecular feature. C, Mean variance explained (ΔR²) across metabolites and proteins under alternative covariate models. Error bars denote standard errors. D, Unique variance explained by self-reported race/ethnicity versus genetic principal components (gPCs) for metabolites. Each point represents one metabolite, with colour indicating the difference in variance explained (ΔR² = Race – gPCs). Labeled metabolites highlight those with the largest discrepancies. E, Unique variance explained by race/ethnicity versus gPCs for proteins, with discordant molecules highlighted. Variance components were estimated using linear regression models, with unique and shared variance derived from block-level decomposition.

We next identified molecules where the contributions of race and ethnicity and ancestry diverged most strongly (Figure 5D–E). In the metabolome, genetic ancestry showed stronger associations with lipid subclasses—including TG 50:3, TG 58:9, TG 58:11, TG 60:12, and N(6)-dimethyl lysine—highlighting the role of genetic background in shaping lipid composition and methylation-related pathways. Conversely, several unannotated metabolite features were more strongly linked to race and ethnicity, pointing to environmental or sociocultural influences not captured by genetic ancestry. In the proteome, ancestry was uniquely associated with the minor apolipoprotein APOF and complement-related proteins (CD300LF, CFHR2), whereas race and ethnicity explained greater variance in immune and inflammatory proteins, including PTGR1, CBLN1, CD33, and IGFBP1. Although the discovery of protein associations may be constrained by the limited number of proteins assayed and the reliance on two cohorts, the metabolite associations—particularly those with genetic ancestry—showed consistency across studies, underscoring their robustness.

Finally, we further dissected the contributions of self-reported race and ethnicity versus genetically inferred ancestry proportions (Supplementary Figure 5). Overall, genetically inferred ancestry explained a greater share of proteomic variance than self-reported race across comparisons. For example, in the Non-Hispanic Black vs. African ancestry and Hispanic vs. American ancestry contrasts, ancestry proportions accounted for substantially more variance, with strong associations for proteins such as APOF, CFHR2, and CD33. Likewise, select proteins displayed stronger associations with genetic ancestry, including IGFBP3 and CFHR2 in the Hispanic vs. American ancestry group. In the Non-Hispanic White vs. European ancestry comparison, ancestry again explained more variance overall, though proteins such as CTSH, BGLAP, and ICAM5 emerged as top genetic contributors. Together, these results highlight that, while correlated, self-reported race and genetic ancestry capture partially distinct biological and social axes of molecular variability.

### Environmentally and Genetic Ancestry Influenced Molecular Signatures and Causal Links to Type 2 Diabetes Risk

To assess the clinical and causal relevance of molecular signatures shaped by environmental and genetic factors (Supplementary Figure 6), we evaluated their associations with incident T2D in meta-analyses across MESA and WHI (Supplementary Tables 4 and 5). Biomarkers were stratified by the domains explaining their variance and tested for both prospective associations and causal contributions.

Environmentally influenced metabolites, including diacylglycerols (DG 36:4), phosphatidylethanolamines (PE 38:6), were consistently associated with increased T2D risk (Figure 6A). Proteins linked to inflammation and vascular function^30–33^, such as ICAM1, ITGB2, and MMP12, showed similar associations (Figure 6B). Genetic ancestry metabolites such as triglyceride species (TG 50:3, TG 52:4, TG 51:3) and proteins including IGFBP3 and IL18R1 showed positive associations with incident T2D, reflecting inherited pathways of metabolic risk. Together, these findings suggest that inherited risk is shaped predominantly by lipid metabolism, adipocyte differentiation, and inflammatory signaling. ^34,35^

**Figure 6.**
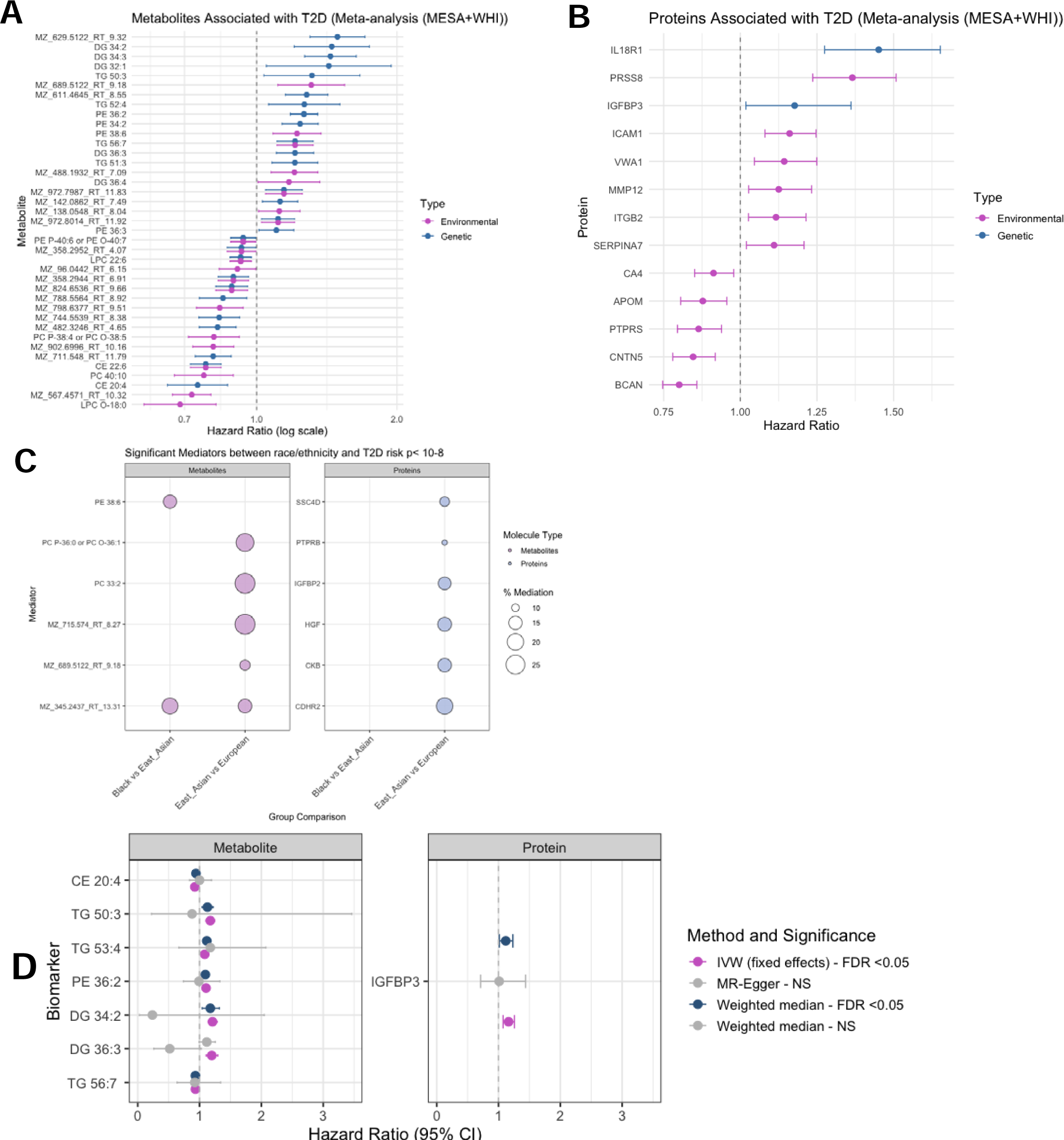
Environmentally and genetically shaped molecular signatures and causal pathways to type 2 diabetes. (A) Prospective associations of metabolites (left) and proteins (right) with incident type 2 diabetes (T2D) in meta-analyses across MESA and WHI. Molecules were classified as predominantly environmentally influenced (pink) or genetically influenced (blue) based on variance decomposition. Points represent hazard ratios (HRs), and horizontal lines indicate 95% confidence intervals. (B) Causal mediation analysis of environmentally influenced molecules in the relationship between race/ethnicity and T2D risk in MESA. Significant mediators (P < 10^-8^) are shown, with circle size denoting the proportion of T2D risk mediated. (C) Two-sample Mendelian randomization of genetically influenced metabolites and proteins, testing their causal effects on T2D. Points represent causal effect estimates (HRs), and horizontal lines denote 95% confidence intervals across three methods: inverse variance weighted (IVW), weighted median, and MR-Egger.

We next investigated causality using complementary inference approaches. Mediation analyses revealed that environmentally driven molecules partially explained population-level disparities in T2D risk (Figure 6C; Supplementary table 6). Several lipid metabolites, including PE 38:6, PC 33:2, and PC O-36:1/PC P-36:0, mediated ∼10–25% of the excess risk observed between East Asian and White groups. Additional unannotated metabolites also emerged as significant mediators, highlighting the contribution of novel, yet uncharacterized signatures. On the proteomic side, IGFBP2, HGF, PTBPB, and SSC4D mediated disparities across East Asian, and White individuals, together with CDHR2 and CKß, implicating immune, growth, and adhesion pathways in environmentally patterned T2D risk.

Finally, to test whether genetic ancestry influenced molecules have a direct causal role in T2D, we applied two-sample Mendelian randomization across metabolites and proteins using IVW, Weighted Median, and MR-Egger approaches (Figure 6D, Supplementary Table 7). Seven lipid species—including CE 20:4, TG 50:3, TG 53:4, PE 36:2, DG 34:2, DG 36:3, and TG 56:7—demonstrated significant associations (FDR < 0.05). The metabolite CE 20:4 was inversely associated with T2D, whereas multiple diacylglycerols and triglycerides conferred higher risk. In the proteome, IGFBP3 emerged as the sole protein with robust causal evidence. Importantly, sensitivity analyses showed no evidence of horizontal pleiotropy (*P >0.05*), supporting the robustness of these findings (Supplementary Table 7). Together, these results reveal complementary causal pathways to T2D: environmentally driven molecules that mediate population-level disparities and genetically determined lipids and proteins that exert direct biological effects.

## DISCUSSION

Biological and lifestyle factors explained the largest molecular variation in plasma metabolites and proteins in two multi-ancestry cohorts. However, self-reported race and ethnicity and genetic ancestry, though more modest, contributed non-redundantly, capturing distinct molecular signatures, including in models additionally adjusted for SES. Distinct biological pathways emerged, with genetic ancestry primarily linked to lipid and apolipoprotein variation, and race and ethnicity enriched for immune and inflammatory processes. Environmentally influenced metabolites—including diacylglycerols (DG 34:2, DG 32:1), phosphatidylethanolamines (PE 36:4, PE 38:4), and lysophosphatidylcholines (LPC 18:0)—as well as proteins involved in inflammation and vascular biology (ICAM1, ITGB2, MMP12, SERPINA7), were consistently associated with higher T2D risk. In contrast, genetically driven triglycerides (e.g., TG 50:3, TG 52:4) and proteins such as IGFBP3 and IL18R1 reflected inherited risk pathways. Causal inference analyses further revealed that specific lipids (e.g., PE 38:6, PC 33:2, PC O-36:1/PC P-36:0) and proteins (IGFBP2, HGF, SSC4D) mediated racial and ethnic disparities in T2D, while Mendelian randomization identified a subset of genetically determined lipids (including CE 20:4, TG 50:3, DG 34:2) and IGFBP3 with putative causal roles.

Our variance decomposition analyses confirmed several well-established associations, including the strong effect of genetic ancestry on lipid levels, indicating that ancestry-driven molecular variation is particularly enriched for lipid and apolipoprotein biology^36–38^. We further replicated known biological and lifestyle associations, such as BMI with triglycerides^20,39,40^, LEP^41^, and FABP4^42^, and lifestyle exposures with omega-3 fatty acids, reflecting dietary intake, and cotinine, a biomarker of smoking. Beyond these confirmatory results, our study uncovered novel patterns whereby self-reported race and ethnicity preferentially captured inflammatory and immune pathways.

This observation aligns with prior work linking systemic and social disadvantage to heightened inflammatory profiles. Race and ethnicity are social constructs that encode differential exposure to structural inequities and lived environments.^39,40^ Individuals from minoritized racial and ethnic groups are more likely to experience socioeconomic disadvantage, residential segregation, and occupational exposures that collectively influence immune and inflammatory biology.^43,44^ These include limited access to education and income, neighborhood environments characterized by higher air pollution, traffic, noise, and reduced green space, as well as chronic psychosocial stress from adverse life experiences and financial insecurity.^45–49^ Occupational factors such as toxic exposures, shift work, and sleep disruption further contribute to heightened inflammatory biology.^44,50^ Collectively, these mechanisms provide biological plausibility for our finding that racial and ethnic identity, beyond genetic ancestry, captures immune- and inflammation-related molecular signatures, highlighting the role of social and environmental determinants in shaping metabolic health disparities.

Our analyses highlight both environmental and genetic pathways through which molecular signatures contribute to T2D risk. Several of the environmentally influenced and genetic ancestry influenced metabolites identified here (DG 34:2, DG 32:1, LPC 18:0, and TG 50:3) have previously been associated with incident T2D in large-scale meta-analyses^4,51^, reinforcing their role in diabetes pathophysiology. Beyond metabolites, our analyses distinguished IGFBP2 and IGFBP3 as representative examples of environmentally versus genetic ancestry influenced proteins. IGFBP2, classified as environmentally influenced, mediated part of the association between self-reported race and ethnicity and T2D, consistent with evidence that relative IGFBP2 deficiency favors visceral adiposity and ectopic lipid storage—established risk factors for T2D. ^52^ By contrast, IGFBP3 was genetic ancestry influenced and supported by Mendelian randomization as having a putative causal role in T2D, reflecting inherited regulation of the IGF axis. Prior studies have similarly identified IGFBP3 as a biomarker linking the absence of ‘favorable adiposity’ with elevated T2D risk and as a predictive marker for incident cardiovascular events.^53^ Together, these findings illustrate how environmentally patterned biology (IGFBP2) and genetic ancestry influenced pathways (IGFBP3) converge to shape diabetes risk through distinct but complementary mechanisms.

Our study advances molecular epidemiology by applying variance decomposition across a wide set of domains including genetic, biological, lifestyle, and social, enabling direct quantification of their unique and shared contributions to proteomic and metabolomic profiles. By jointly considering genetic ancestry and self-reported race and ethnicity, we illustrate how inherited and environmental forces converge to shape T2D risk across populations. Importantly, racial identity is closely intertwined with diet, socioeconomic status (SES), and other contextual exposures. These dimensions are not independent: for example, dietary patterns and SES may act as upstream mediators of observed race and ethnicity associations with T2D risk. Thus, part of the total effect attributed to race and ethnicity likely reflects modifiable social and environmental pathways. Future work applying causal mediation frameworks could clarify these relationships and identify the most actionable intervention targets. Equally important, expanding multi-omic profiling in larger and more diverse populations, and translating molecular signatures into prevention and treatment strategies, will be critical to ensure that precision medicine advances health equity.

Several limitations warrant consideration. First, despite extensive covariate adjustment, residual confounding from unmeasured environmental exposures cannot be excluded. Genetic ancestry is often strongly correlated with social and environmental factors, making it difficult to fully disentangle their independent effects. Consequently, molecular features classified as genetic ancestry–influenced or environmentally influenced may partially capture overlapping domains. Nonetheless, our multivariable modeling approach, which incorporated both biological and socioeconomic covariates, was designed to minimize this overlap and isolate the predominant sources of variability as effectively as possible. Second, our findings are derived from two U.S.-based cohorts (MESA and WHI) and may not generalize to all populations. Third, self-reported race and ethnicity is a dynamic construct and can vary across place and time. Finally, the modest sample size limited our power to detect subtler effects, meaning that some associations may have gone undetected.

These findings have important implications for health equity. By identifying molecular signatures that link sociocultural exposures—such as systemic disadvantage, chronic stress, and environmental conditions—to T2D risk, we provide biological evidence of pathways through which social determinants shape health disparities. Together, these insights emphasize that disentangling social and biological contributions is critical to understanding and ultimately mitigating disparities in metabolic disease.

Finally, our results set the stage for translational research. Environmentally influenced biomarkers may serve as modifiable targets for intervention, while genetic ancestry influenced pathways can inform risk stratification and mechanistic studies. Integrating both perspectives into precision medicine approaches will not only improve prediction of T2D risk across diverse populations but also ensure that molecular insights are leveraged to reduce, rather than exacerbate, health inequities.

## DECLARATION OF INTERESTS

SJC reports a family member employed by Depuy-Synthes. LMR is a consultant for the TOPMed Administrative Coordinating Center (through Westat).

## Data Availability

All data used in this study were obtained through the Trans-Omics for Precision Medicine (TOPMed) program, which integrates data from multiple cohorts including MESA and WHI. These data are available through controlled-access policies of TOPMed via dbGaP. Processed data generated in this study are available from the corresponding author upon reasonable request.

## ACKNOWLEDGEMENTS

NW, PAH, AB, YB-W, KDT, LMR, CK, JD, JIR, CTL, JBM, and AKM are supported by NIDDK UM1 DK078616. MS-G is supported by NIDDK UM1 DK078616 and NIDDK K99 DK139461. SJC is supported by the American Diabetes Association (7-21-JDFM-005). A complete list of NHLBI Trans-Omics for Precision Medicine (TOPMed) Consortium authors and their affiliations is provided in the Supplementary Material.

## TOPMed Acknowledgments

Molecular data for the Trans-Omics in Precision Medicine (TOPMed) program was supported by the National Heart, Lung and Blood Institute (NHLBI). See the TOPMed Omics Support Table below for study specific omics support information. Core support including centralized genomic read mapping and genotype calling, along with variant quality metrics and filtering were provided by the TOPMed Informatics Research Center (3R01HL-117626-02S1; contract HHSN268201800002I). Core support including phenotype harmonization, data management, sample-identity QC, and general program coordination were provided by the TOPMed Data Coordinating Center (R01HL-120393; U01HL-120393; contract HHSN268201800001I). Freeze 1RNA TOPMed cis-eQTL results were generated in a collaboration between the TOPMed Informatics Research Center, TOPMed Multi-Omics working group, and the TOPMed parent studies contributing RNA-seq and distributed to TOPMed investigators. We gratefully acknowledge the studies and participants who provided biological samples and data for TOPMed.

Phase 1 TOPMed metabQTL results were generated in a collaboration between the TOPMed Metabolomics Working Group, the TOPMed Informatics Research Center, and the TOPMed parent studies contributing metabolite data and distributed to TOPMed investigators.

## Study Acknowledgements

MESA and the MESA SHARe project are conducted and supported by the National Heart, Lung, and Blood Institute (NHLBI) in collaboration with MESA investigators. Support for MESA is provided by contracts 75N92025D00022, 75N92020D00001, HHSN268201500003I, N01-HC-95159, 75N92025D00026, 75N92020D00005, N01-HC-95160, 75N92020D00002, N01-HC-95161, 75N92025D00024, 75N92020D00003, N01-HC-95162, 75N92025D00027, 75N92020D00006, N01-HC-95163, 75N92025D00025, 75N92020D00004, N01-HC-95164, 75N92025D00028, 75N92020D00007, N01-HC-95165, N01-HC-95166, N01-HC-95167, N01-HC-95168, N01-HC-95169, UL1-TR-000040, UL1-TR-001079, UL1-TR-001420, UL1TR001881, DK063491, and R01HL105756. The authors thank the MESA participants and the MESA investigators and staff for their valuable contributions. A full list of participating MESA investigators and institutions can be found at http://www.mesa-nhlbi.org. The WHI program is funded by the National Heart, Lung, and Blood Institute, National Institutes of Health, U.S. Department of Health and Human Services through contracts 75N92021D00001, 75N92021D00002, 75N92021D00003, 75N92021D00004, 75N92021D00005.

## Notes

### Author Declarations

The Institutional Review Board of Massachusetts General Hospital gave ethical approval for this work.

