## Supplementary Figures for "Dissecting Genetic and Environmental Determinants of Plasma Molecular Signatures and Their Link to Type 2 Diabetes Risk"

### Slide 1
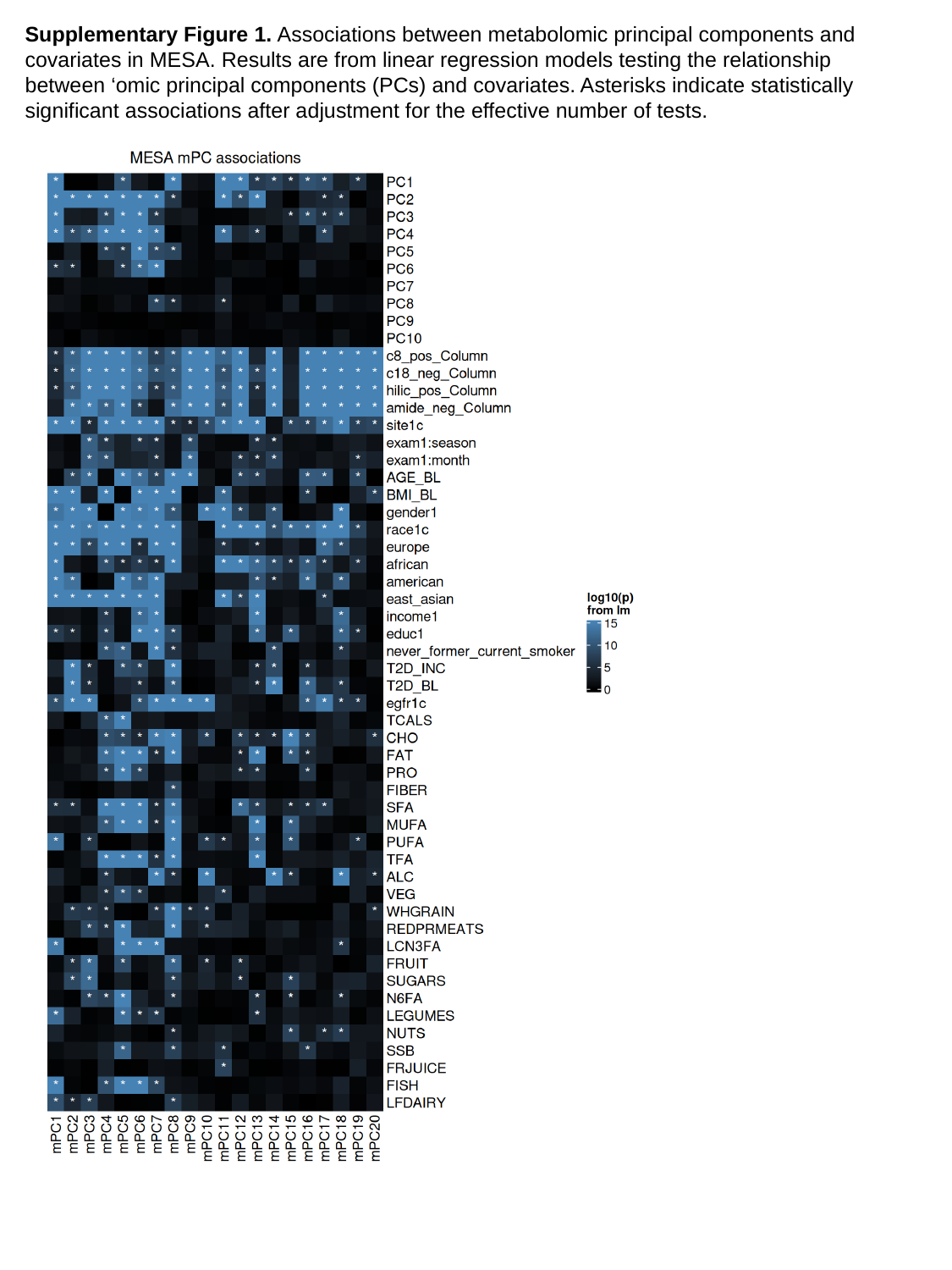

Supplementary Figure 1. Associations between metabolomic principal components and covariates in MESA. Results are from linear regression models testing the relationship between ‘omic principal components (PCs) and covariates. Asterisks indicate statistically significant associations after adjustment for the effective number of tests.

### Slide 2
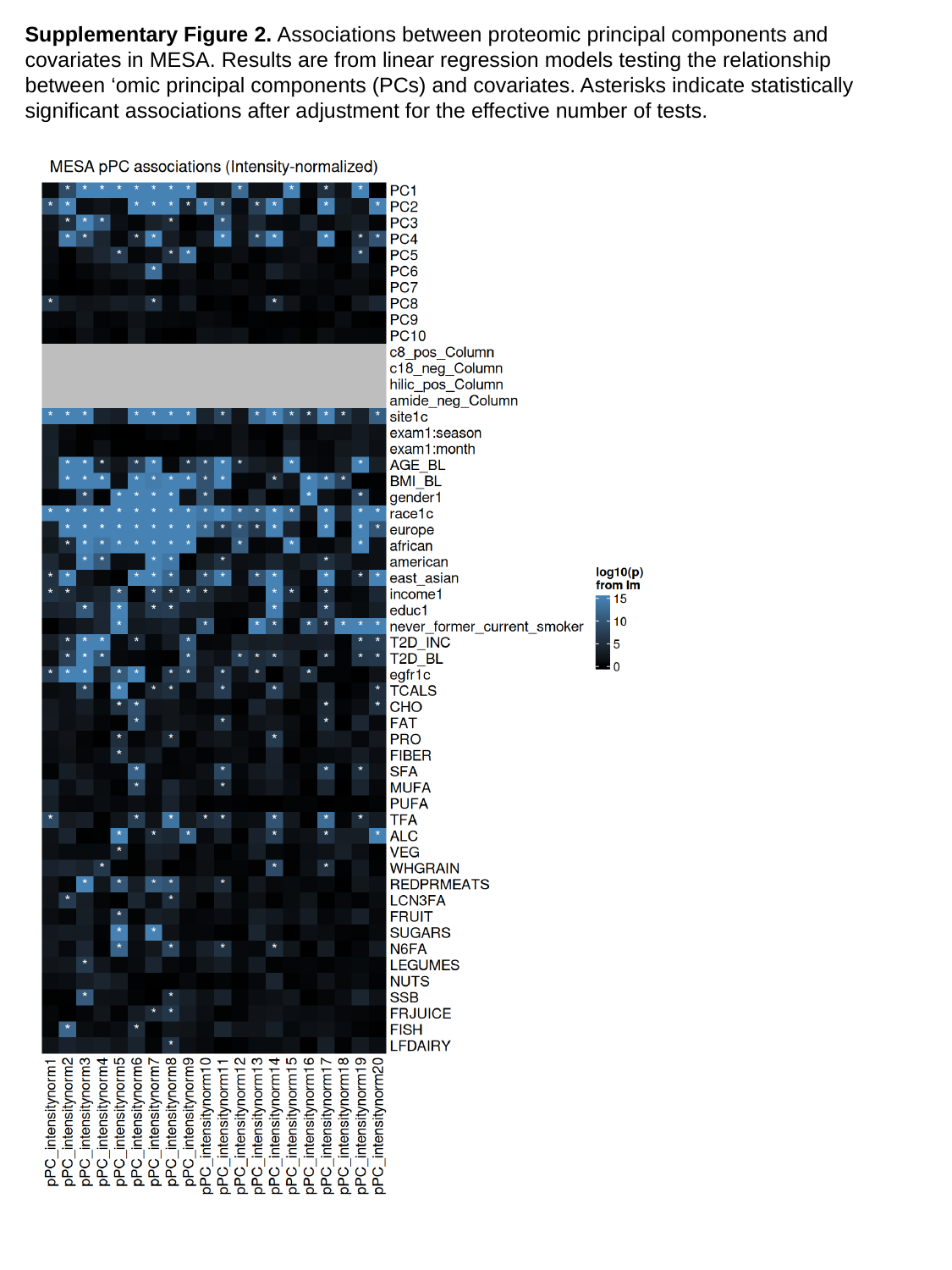

Supplementary Figure 2. Associations between proteomic principal components and covariates in MESA. Results are from linear regression models testing the relationship between ‘omic principal components (PCs) and covariates. Asterisks indicate statistically significant associations after adjustment for the effective number of tests.

### Slide 3
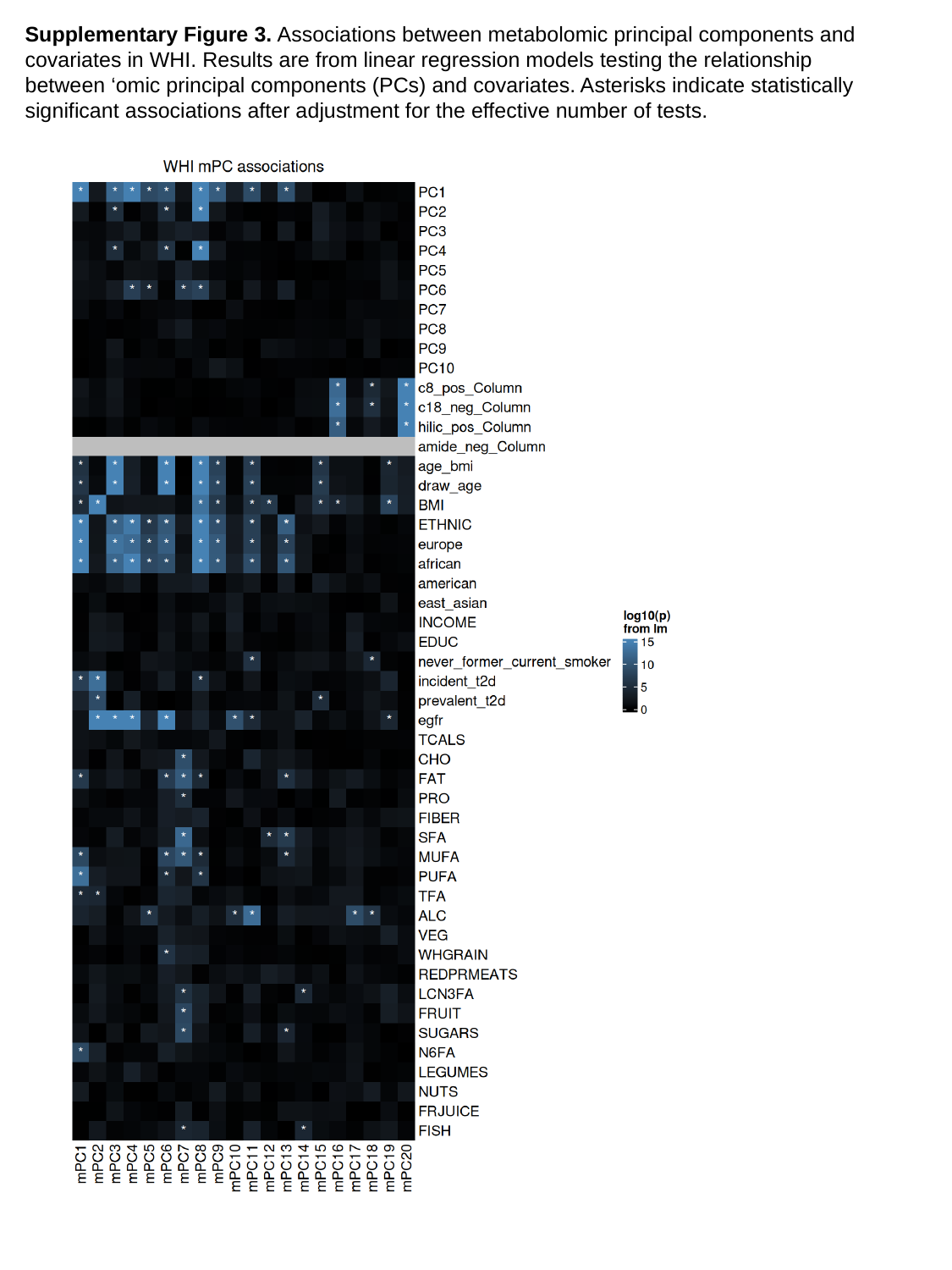

Supplementary Figure 3. Associations between metabolomic principal components and covariates in WHI. Results are from linear regression models testing the relationship between ‘omic principal components (PCs) and covariates. Asterisks indicate statistically significant associations after adjustment for the effective number of tests.

### Slide 4
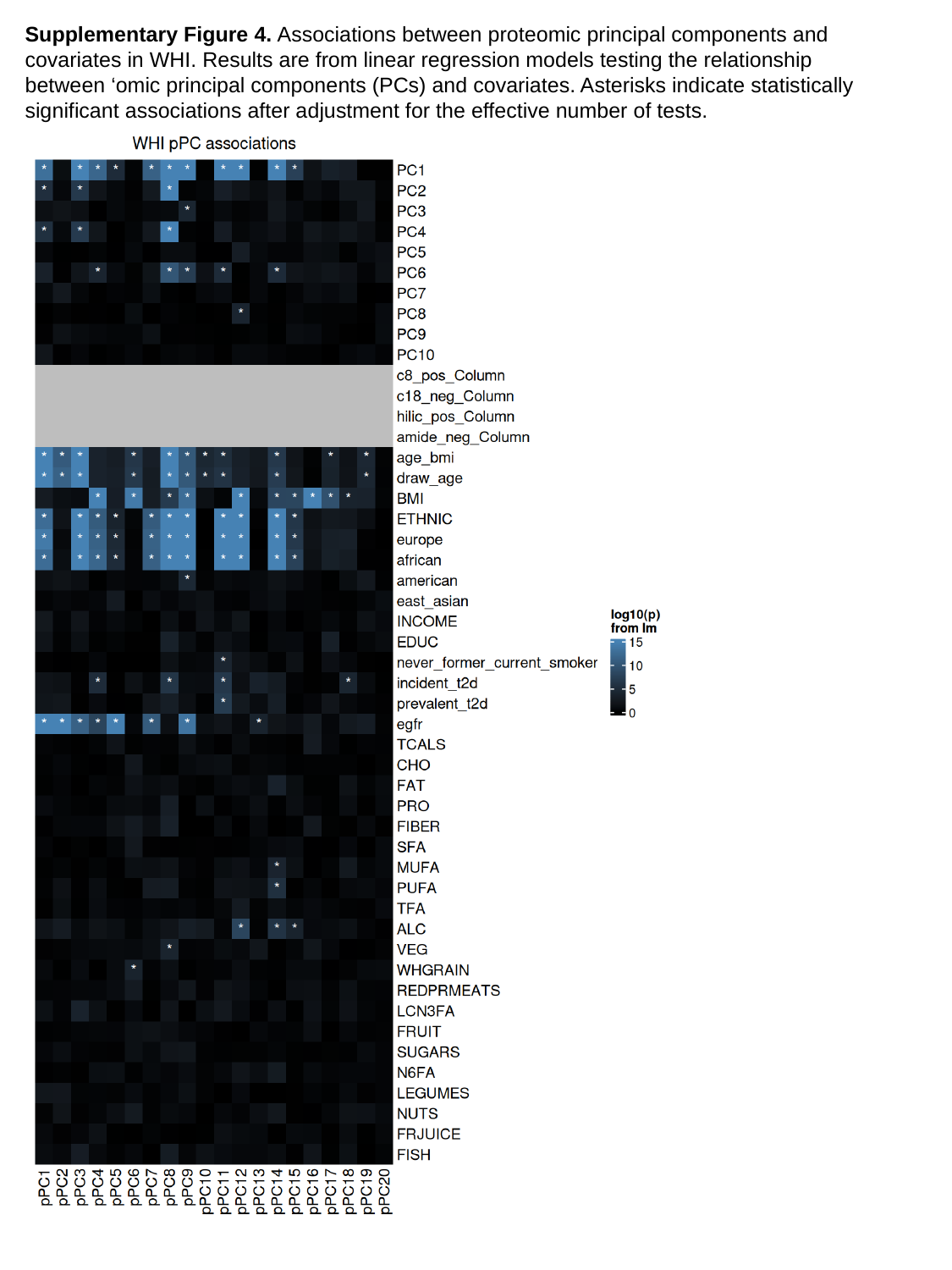

Supplementary Figure 4. Associations between proteomic principal components and covariates in WHI. Results are from linear regression models testing the relationship between ‘omic principal components (PCs) and covariates. Asterisks indicate statistically significant associations after adjustment for the effective number of tests.

### Slide 5
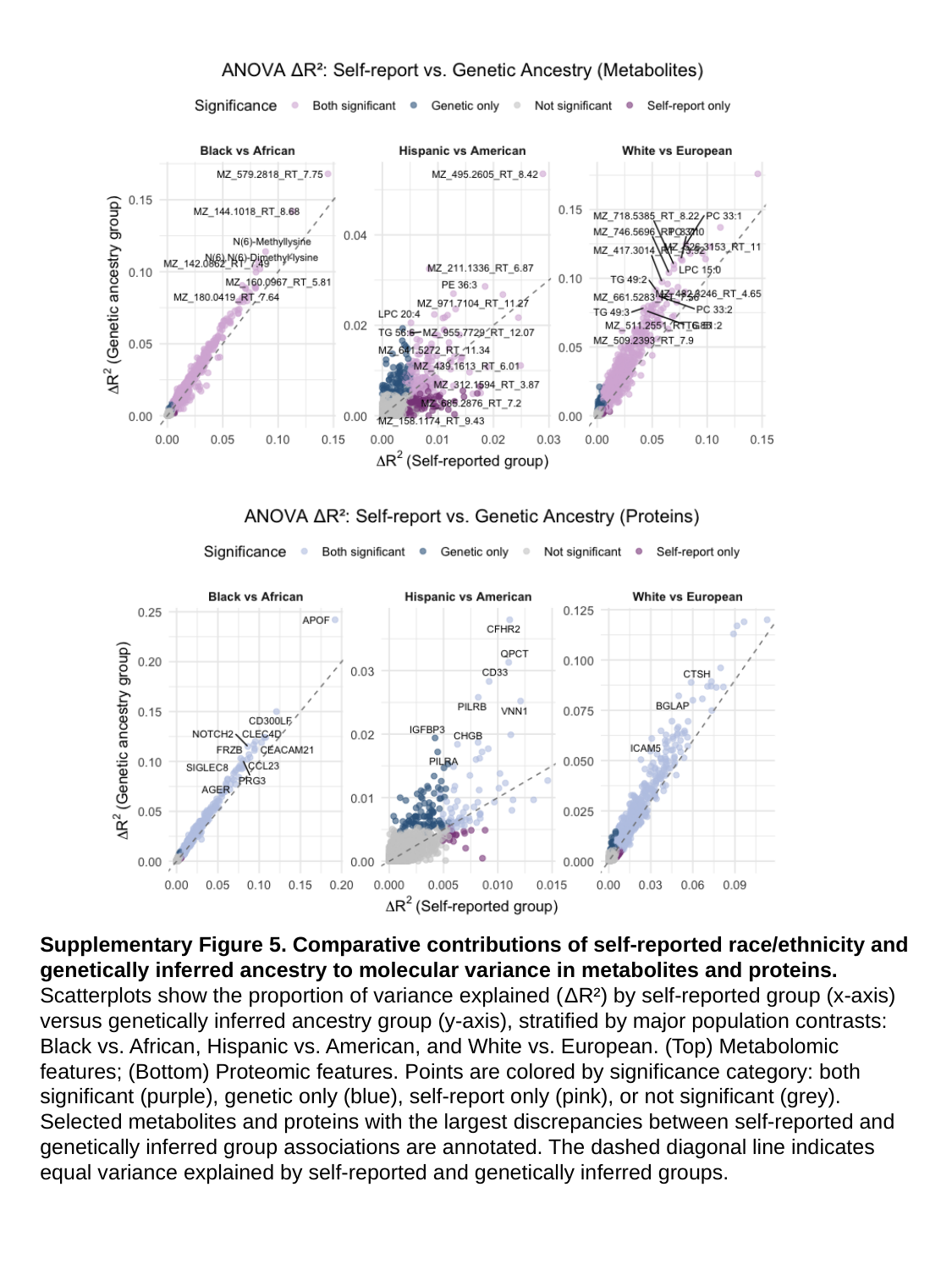

Supplementary Figure 5. Comparative contributions of self-reported race/ethnicity and genetically inferred ancestry to molecular variance in metabolites and proteins.
Scatterplots show the proportion of variance explained (ΔR²) by self-reported group (x-axis) versus genetically inferred ancestry group (y-axis), stratified by major population contrasts: Black vs. African, Hispanic vs. American, and White vs. European. (Top) Metabolomic features; (Bottom) Proteomic features. Points are colored by significance category: both significant (purple), genetic only (blue), self-report only (pink), or not significant (grey). Selected metabolites and proteins with the largest discrepancies between self-reported and genetically inferred group associations are annotated. The dashed diagonal line indicates equal variance explained by self-reported and genetically inferred groups.

### Slide 6
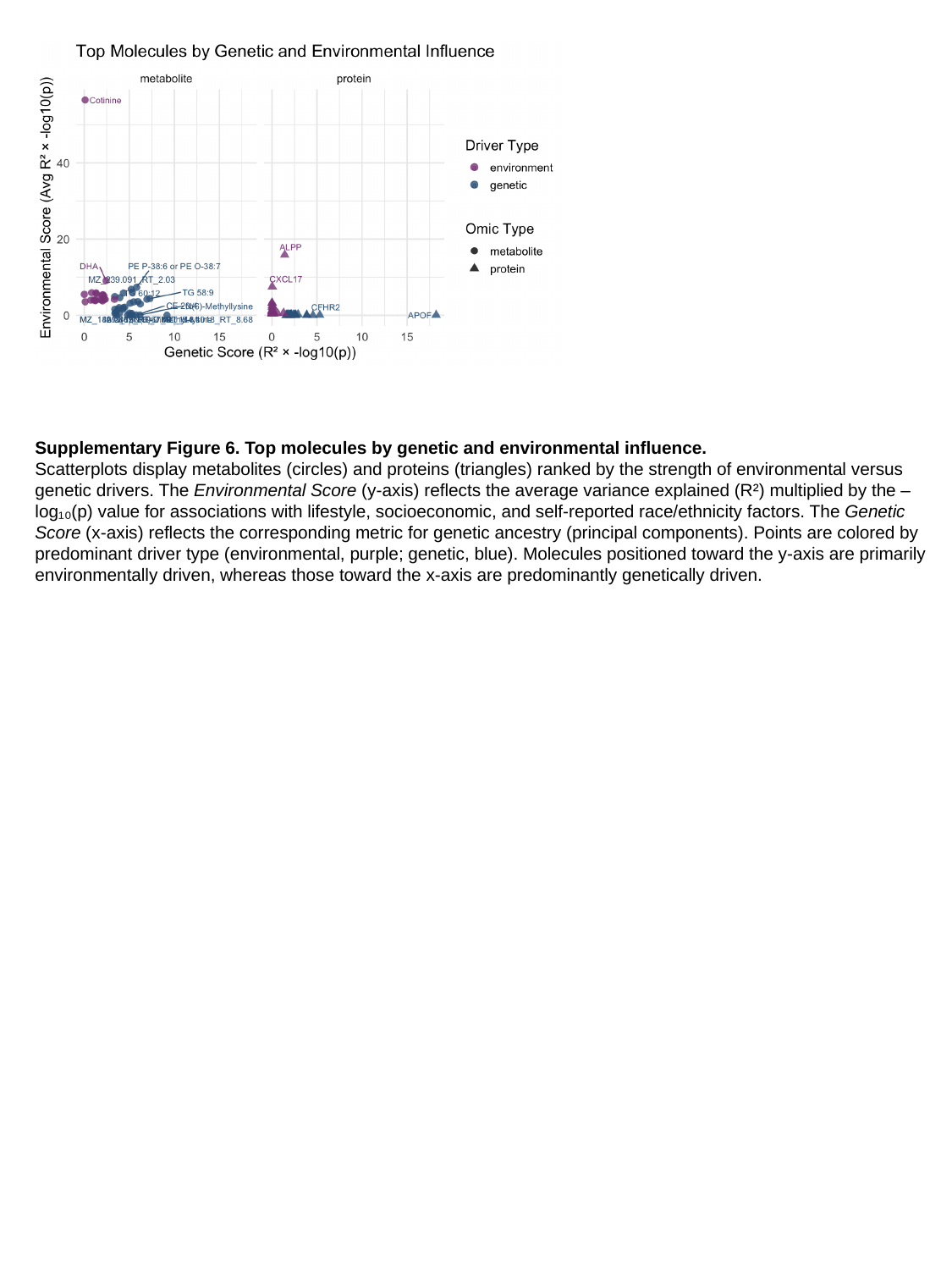

Supplementary Figure 6. Top molecules by genetic and environmental influence.
Scatterplots display metabolites (circles) and proteins (triangles) ranked by the strength of environmental versus genetic drivers. The Environmental Score (y-axis) reflects the average variance explained (R²) multiplied by the –log₁₀(p) value for associations with lifestyle, socioeconomic, and self-reported race/ethnicity factors. The Genetic Score (x-axis) reflects the corresponding metric for genetic ancestry (principal components). Points are colored by predominant driver type (environmental, purple; genetic, blue). Molecules positioned toward the y-axis are primarily environmentally driven, whereas those toward the x-axis are predominantly genetically driven.

### Slide 7
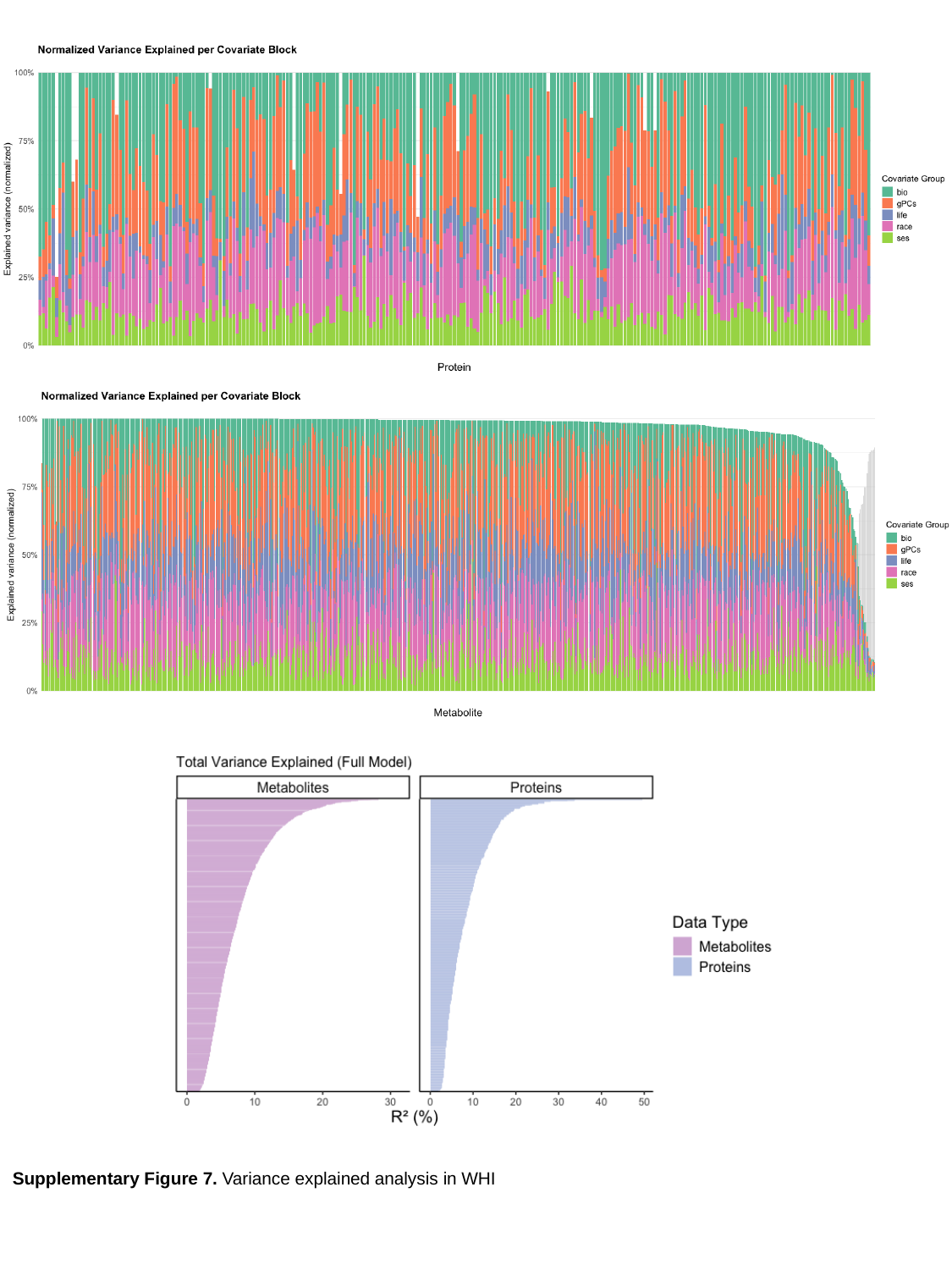

Supplementary Figure 7. Variance explained analysis in WHI

### Slide 8
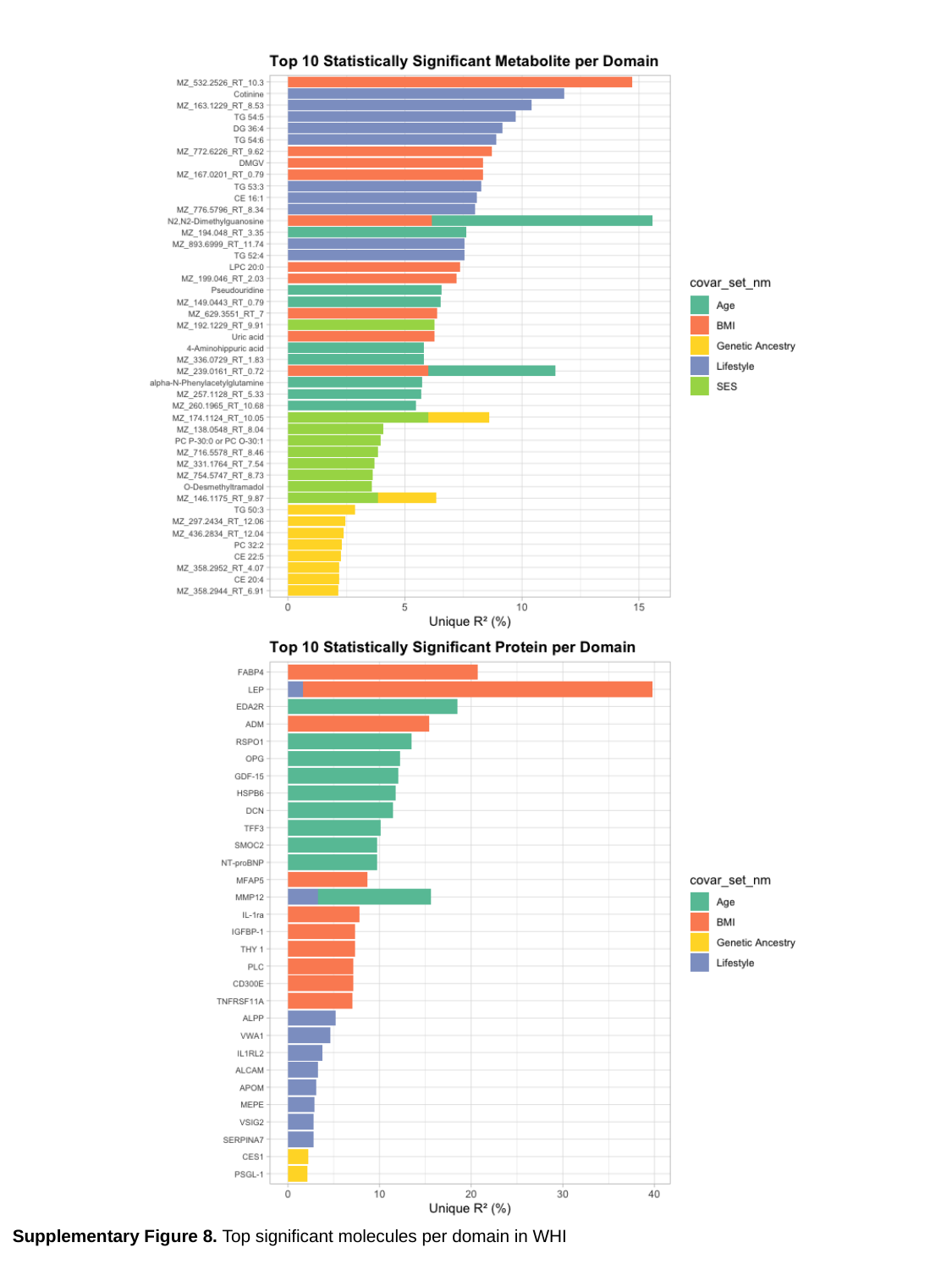

Supplementary Figure 8. Top significant molecules per domain in WHI

### Slide 9
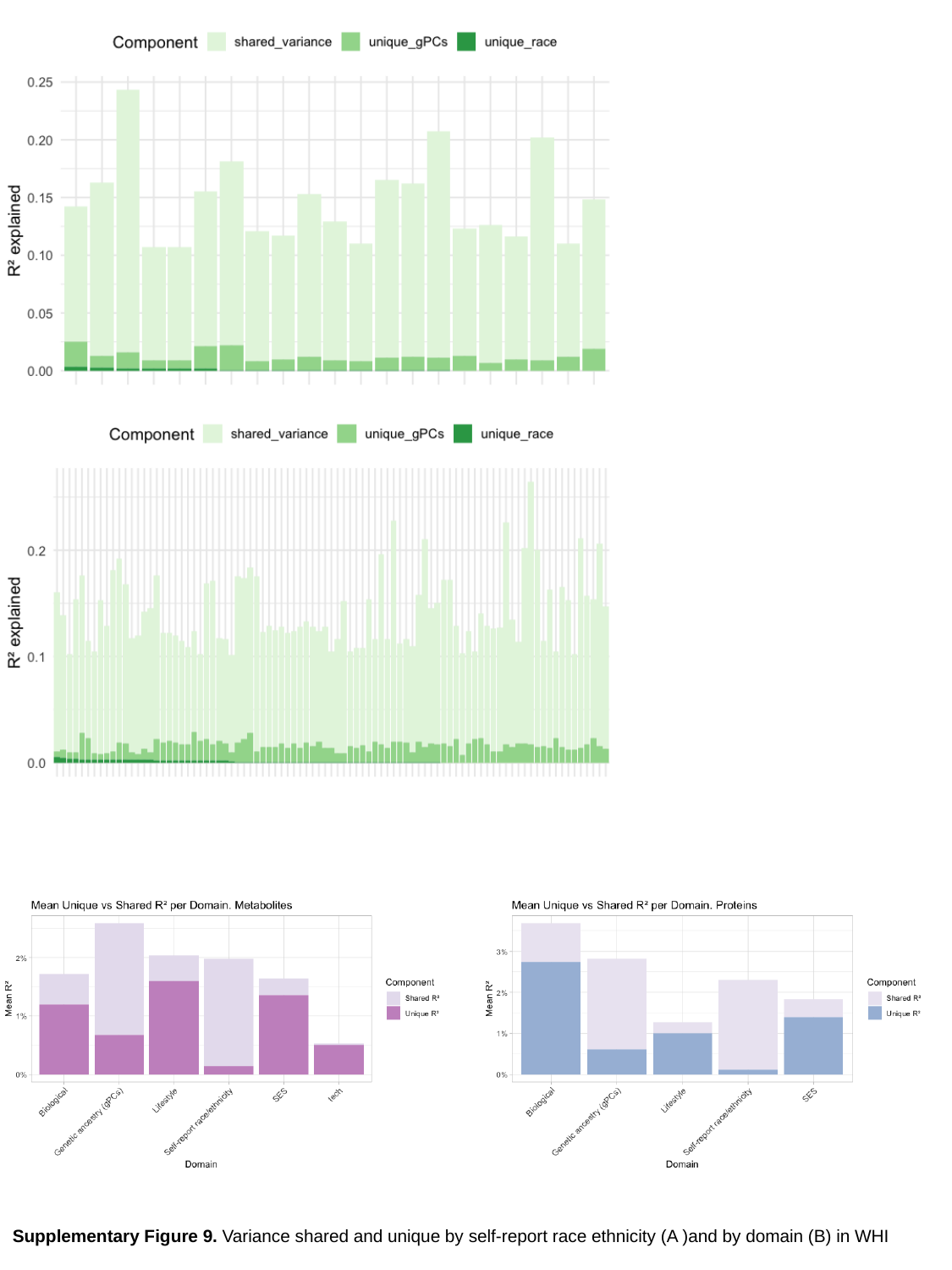

Supplementary Figure 9. Variance shared and unique by self-report race ethnicity (A )and by domain (B) in WHI

### Slide 10
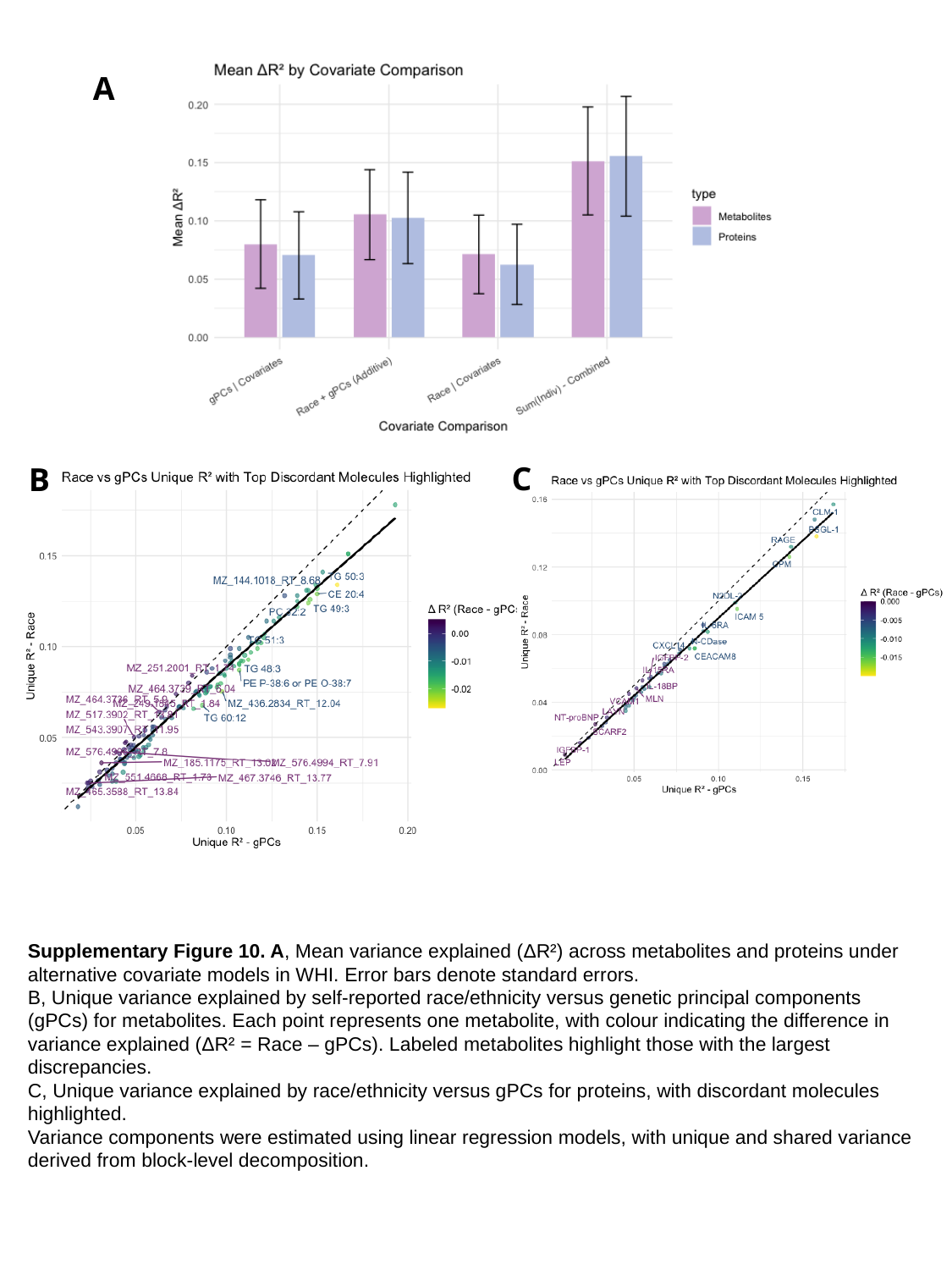

A
C
B
Supplementary Figure 10. A, Mean variance explained (ΔR²) across metabolites and proteins under alternative covariate models in WHI. Error bars denote standard errors.
B, Unique variance explained by self-reported race/ethnicity versus genetic principal components (gPCs) for metabolites. Each point represents one metabolite, with colour indicating the difference in variance explained (ΔR² = Race – gPCs). Labeled metabolites highlight those with the largest discrepancies.
C, Unique variance explained by race/ethnicity versus gPCs for proteins, with discordant molecules highlighted.
Variance components were estimated using linear regression models, with unique and shared variance derived from block-level decomposition.

### Slide 11
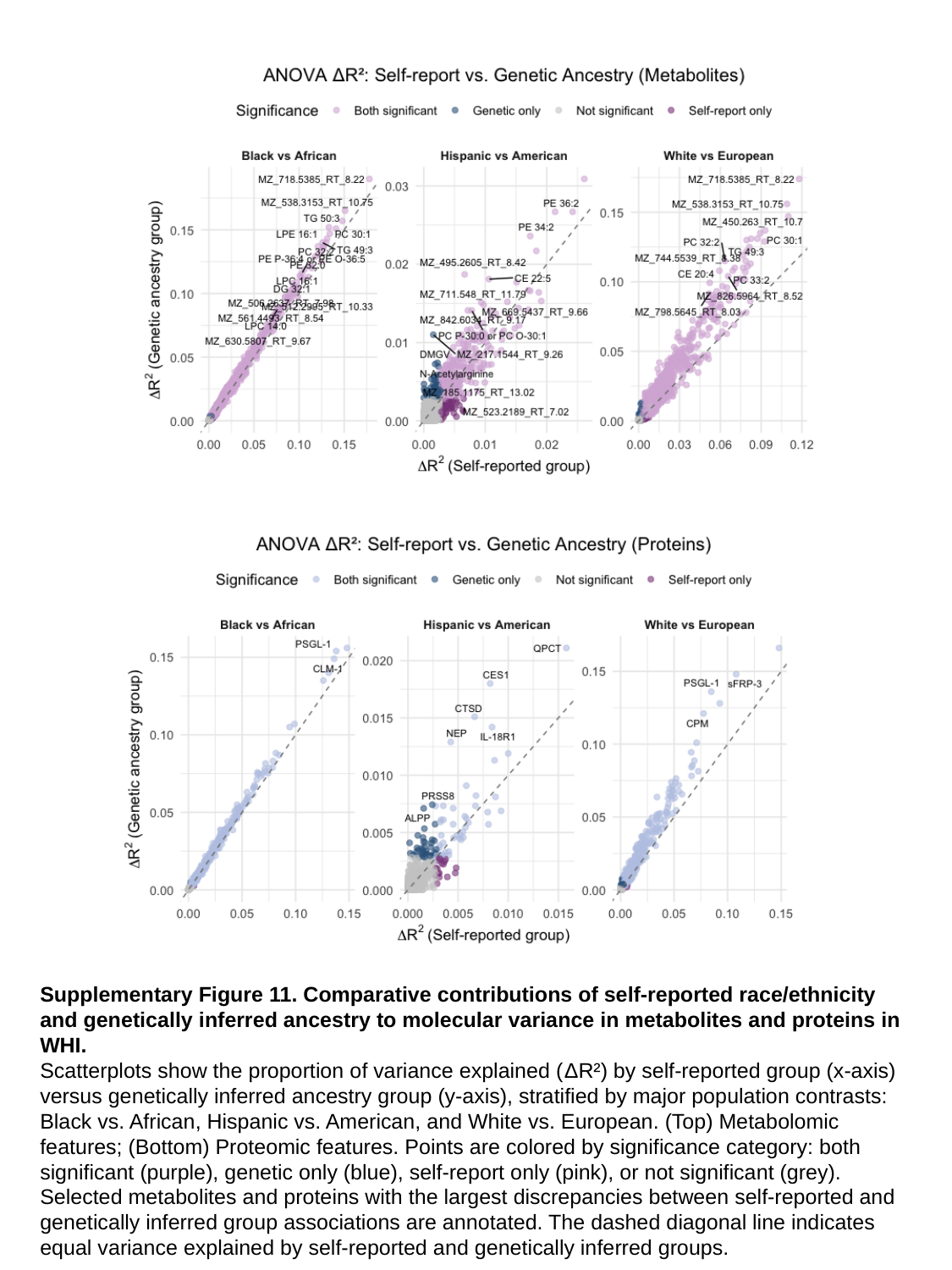

Supplementary Figure 11. Comparative contributions of self-reported race/ethnicity and genetically inferred ancestry to molecular variance in metabolites and proteins in WHI.
Scatterplots show the proportion of variance explained (ΔR²) by self-reported group (x-axis) versus genetically inferred ancestry group (y-axis), stratified by major population contrasts: Black vs. African, Hispanic vs. American, and White vs. European. (Top) Metabolomic features; (Bottom) Proteomic features. Points are colored by significance category: both significant (purple), genetic only (blue), self-report only (pink), or not significant (grey). Selected metabolites and proteins with the largest discrepancies between self-reported and genetically inferred group associations are annotated. The dashed diagonal line indicates equal variance explained by self-reported and genetically inferred groups.

### Slide 12
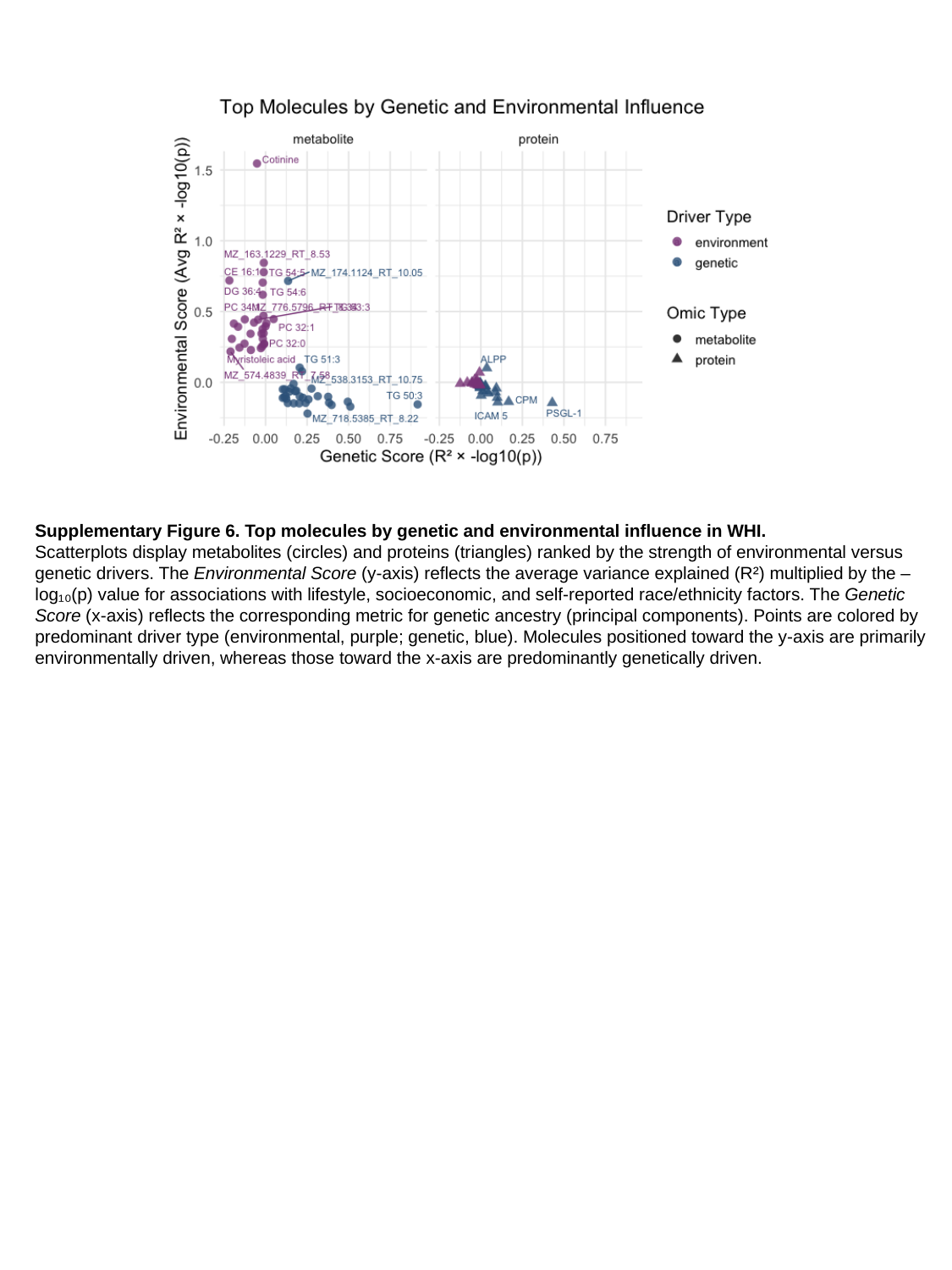

Supplementary Figure 6. Top molecules by genetic and environmental influence in WHI.
Scatterplots display metabolites (circles) and proteins (triangles) ranked by the strength of environmental versus genetic drivers. The Environmental Score (y-axis) reflects the average variance explained (R²) multiplied by the –log₁₀(p) value for associations with lifestyle, socioeconomic, and self-reported race/ethnicity factors. The Genetic Score (x-axis) reflects the corresponding metric for genetic ancestry (principal components). Points are colored by predominant driver type (environmental, purple; genetic, blue). Molecules positioned toward the y-axis are primarily environmentally driven, whereas those toward the x-axis are predominantly genetically driven.
